# Prognostic predictions in psychosis: exploring the complementary role of machine learning models

**DOI:** 10.1101/2025.01.30.25321382

**Authors:** Violet van Dee, Seyed Mostafa Kia, Caterina Fregosi, Wilma E. Swildens, Anne Alkema, Albert Batalla, Coen van den Berg, Danko Coric, Edwin van Dellen, Lotte G. Dijkstra, Arthur van den Doel, Livia S. Dominicus, John Enterman, Frank L. Gerritse, Marte Z. van der Horst, Fedor van Houwelingen, Charlotte S. Koch, Lisanne E.M. Koomen, Marjan Kromkamp, Michelle Lancee, Brian E. Mouthaan, Diane F. van Rappard, Eline J. Regeer, Raymond W.J. Salet, Metten Somers, Jorgen Straalman, Marjolein H.T. de Vette, Judith Voogt, Inge Winter-van Rossum, René S. Kahn, Wiepke Cahn, Hugo G. Schnack

**Affiliations:** Department of Psychiatry, UMC Utrecht Brain Center, University Medical Center, Utrecht, the Netherlands; Department of Psychiatry, St Antonius Hospital, Utrecht, The Netherlands; Donders Institute for Brain, Cognition, and Behaviour, Radboud University, Nijmegen, the Netherlands; Department of Cognitive Science and Artificial Intelligence, Tilburg University, Tilburg, the Netherlands; Department of Informatics, System and Communication, University of Milano-Bicocca, Milan, Italy; Altrecht Institute for Mental Health Care, Utrecht, The Netherlands; Inholland University of Applied Sciences, Amsterdam, The Netherlands; GGZ Delfland, Delft, The Netherlands; Department of Neurology, UZ Brussel and Vrije Universiteit Brussel, Brussels, Belgium; Parnassia Group for Mental Health Care, The Hague, The Netherlands; Department of Psychiatry, Tergooi MC, Hilversum, The Netherlands; Mediant Mental Health Care, Enschede, The Netherlands; BuurtzorgT Mental Health Care, Utrecht, the Netherlands; Department of Psychiatry, Icahn School of Medicine at Mount Sinai, New York City, USA; Institute for Language Sciences, Utrecht University, Utrecht, the Netherlands

## Abstract

**BACKGROUND:** Predicting outcomes in schizophrenia spectrum disorders is challenging due to the variability of individual trajectories. While machine learning (ML) shows promise in outcome prediction, is has not yet been integrated into clinical practice. Understanding how ML models (MLMs) can complement psychiatrists’ predictions and bridge the gap between MLM capabilities and practical use is key.

**OBJECTIVE:** This study aims to compare the performance of psychiatrists and MLMs in predicting short-term symptomatic and functional remission in patients with first-episode psychosis and explore whether MLMs can improve psychiatrists’ prognostic accuracy.

**METHOD:** Twenty-four psychiatrists predicted symptomatic and functional remission probabilities based on written baseline information from 66 patients in the OPTiMiSE trial. ML-generated predictions were then shared with psychiatrists, allowing them to adjust their estimates. A questionnaire assessed trust in MLMs, perceived information gaps, and psychiatrists’ self-assessed predictive accuracy, which was compared to actual accuracy.

**FINDINGS:** The predictive accuracy of the MLM was comparable to that of psychiatrists for symptomatic remission (MLM: 0.50, psychiatrists: 0.52) and functional remission (MLM: 0.72, psychiatrists: 0.79). Interrater agreement was low but comparable for psychiatrists and the MLM. Although the MLM did not improve overall predictive accuracy, it showed potential in aiding psychiatrists with difficult-to-predict cases. However, psychiatrists struggled to recognize when to rely on the model’s output and we were unable to determine a clear pattern in these cases based on their characteristics. Psychiatrists could not reliably estimate their predictive accuracy. Psychiatrists expressed moderate to high trust in MLMs for prognostic prediction, but highlighted concerns about the lack of transparency and interpretability of model outputs.

**CONCLUSIONS:** MLMs are a promising tool for supporting psychiatric decision-making, particularly in challenging cases. However, their potential remains underutilized due to limitations in predictive accuracy and a lack of clarity in how predictions are generated. Addressing these issues is essential to build trust and foster integration into clinical practice.

**CLINICAL IMPLICATIONS:** MLMs are best suited as supplementary tools, providing a second opinion while psychiatrists retain decision-making autonomy. Integrating predictions from both sources may help reduce individual biases and improve accuracy. This approach leverages the strengths of MLMs without compromising clinical responsibility.

**SUMMARY BOX:** *What is already known on this topic:* While machine learning models (MLMs) show promise in predicting outcomes in psychotic disorders, they have yet to be integrated into clinical practice. Evidence on the predictive accuracy of psychiatrists for these disorders is limited, with only two small studies published before 1990 suggesting moderate accuracy. Comparisons of MLMs and psychiatrists in this context have not been previously conducted.

*What this study adds:* This is the first study to compare the predictive accuracy of psychiatrists with that of an MLM for psychotic disorders and to assess whether an MLM can enhance psychiatrists’ performance. It highlights that while MLMs do not improve overall accuracy, they may support psychiatrists in difficult cases. Insights into psychiatrists’ trust in MLMs and the challenges of implementing these models are also provided.

*How this study might affect research, practice, or policy:* The findings emphasize the need for advancements in MLM accuracy, interpretability, and strategies to identify cases where MLMs are most beneficial. These improvements could foster effective integration of MLMs as supplementary tools in clinical practice, aiding psychiatrists in decision-making while maintaining their autonomy.

## BACKGROUND

Schizophrenia spectrum disorders have highly variable outcomes, making accurate and individualized outcome prediction - essential to personalized care - a challenging task. The integration of machine learning models (MLMs) into clinical psychiatry presents a promising opportunity to enhance precision in predicting these outcomes. MLMs can detect patterns and statistical dependencies in large datasets, which can be used to support data-driven clinical decision-making. Over the past two decades, numerous studies utilizing MLMs for outcome prediction in SSD have emerged.^1-3^ However, many of these studies are limited by small sample sizes and a lack external validation, weakening the robustness of their findings.^1 4 5^

Despite these limitations, ongoing advances in algorithmic predictive power signal that the time has come to bridge the gap between MLM capabilities and clinical practice. Currently, no MLMs for psychiatric disorders, including SSDs, have been integrated into clinical practice yet.^6 7^ Establishing their clinical value requires comparison to clinicians’ predictions, particularly regarding accuracy, interrater reliability, and potential to assist or augment traditional assessments.

Current studies mainly assess MLMs by comparing their predictive accuracy to random chance. For clinical relevance, however, MLM performance should be compared to clinicians’ accuracy. If an MLM performs at least as well as the psychiatrist, it could potentially assist or replace the psychiatrist for this task. Little data exists on the accuracy and interrater reliability of psychiatrists’ outcome predictions.^8^

Psychiatrists predictions rely on demographic and clinical information, DSM 5 classifications, personal clinical experience and ‘skilled’ intuition.^9 10^ The multifactorial etiology of psychiatric symptoms, the lack of objective biomarkers, incomplete information, questionable validity of psychiatric classifications, and susceptibility of clinical impression to bias may contribute to low accuracy and low inter-rater agreement of psychiatrists in clinical predictions.^9 11 12^ Regarding schizophrenia spectrum disorders, only two small studies published before 1990 have investigated the prognostic accuracy of outcome prediction by clinicians.^13 14^ In the first study, psychiatrists’ predictions on both clinical and functional outcome parameters after 1 year scarcely outperformed chance statements.^13^ The second study reported ‘acceptable agreement’ between predictions and true outcome, particularly for clinical outcomes (‘length of psychotic episode’ and ‘time spent in hospital’) compared to functional outcomes (‘occupational capacity’ and ‘functioning in family’).^14^ Discrepancies in predictive accuracy between studies might stem from differences in defining the criteria for correct predictions between studies. Interrater reliability of psychiatrists in SSD remains unexamined.

## OBJECTIVE

In our study, psychiatrists and MLM predicted individual chance of symptomatic and functional remission at 10 weeks for 66 patients with first episode psychosis, solely based on the collected baseline information. With the generated predictions, we aimed to 1) compare the predictive performance of psychiatrists versus MLMs, 2) explore whether MLM helped psychiatrists to enhance the accuracy of their prognosis predictions, 3) assess the accuracy of psychiatrists in estimating their own predictions, and 4) evaluate the trust of psychiatrists in MLMs.

## METHODS

### STUDY DESIGN

This study employed a longitudinal cohort design to investigate the predictive accuracy of both psychiatrists and a MLM in predicting symptomatic and functional remission in people with first episode psychosis. This study was preregistered on AsPredicted.org (protocol #132306).

### DATA COLLECTION

#### OPTiMiSE dataset

In this study, data from the OPTiMiSE trial were utilized (trial identifier number NCT01248195).^15^ The primary objective of the OPTiMiSE trial was to establish a treatment algorithm for individuals with first episode schizophrenia. The first phase involved 446 patients undergoing amisulpride treatment for four weeks. This phase was completed by 371 patients. Patients meeting the symptom severity component of the consensus criteria for symptomatic remission of the Remission in Schizophrenia Working Group (RSWG), automatically completed the study.^16^ Subsequently, the 93 patients not in remission proceeded to the second phase, where they were randomly assigned to either continue amisulpride or switch to olanzapine for an additional six weeks in a double-blind fashion.

For the current study, data from the 66 patients that completed the second phase with complete Positive And Negative Symptom Scale (PANSS) records were used. Characteristics of these patients are available in Supplement 1. For all included patients the ‘true outcome’ at 10 weeks follow-up was established. Symptomatic remission was defined according to the symptom severity component of the RSWG-criteria ^16^. Functional remission was defined as a score of 71 points and higher on the Personal and Social Performance scale (PSP), because a score between 71-100 refers to only mild difficulties.^17^

#### Machine learning model

The psychosis prognosis prediction model proposed by van Opstal et al.^18^ was employed in this study. This model, based on a recurrent neural network architecture, accommodates multi-model data from diverse sources and can simultaneously predict multiple outcome measures. The model was trained using the OPTiMiSE dataset and used to predict symptomatic and functional outcomes at 10 weeks for those patients not used for training.^18^ Available patient information collected at baseline consisted of demographic, lifestyle, somatic and diagnostic information and data of several questionnaires used for symptom screening and scoring (Table 1).

**Table 1.** Patient information and measurements.

| Type | Number of Features | Features |
| --- | --- | --- |
| Baseline information available to psychiatrists and MLM for making predictions |  |  |
| Demographic | 20 | Age (con), Sex (bin), Race (cat), Immigration status (bin), Marital status (bin), Divorce status (bin), Occupation status (bin), Occupation type (cat), Previous occupation status (bin), Previous occupation type (cat), Father’s occupation (cat), Mother’s occupation (cat), Years of education (con), Highest education level (cat), Father’s highest degree (cat), Mother’s highest degree (cat), Living status (bin), Dwelling (cat), Income source (cat), Living environment (cat) |
| Diagnostic | 7 | DSM-IV classification (cat), Duration of the current psychotic episode (con), Current psychiatric treatment (cat), Psychosocial interventions status (bin), Estimated prognosis (cat), Hospitalization status (bin) |
| Lifestyle | 7 | Recreational drugs history (bin), Recreational drugs since last visit (bin), Caffeine drinks per day (con), Last caffeine drink (cat), Drink Alcohol (bin), Alcoholic drinks in the last year (cat), Smoking status (bin) |
| Somatic | 11 | Height (con), Weight (con), Waist (con), Hip (con), BMI (con), Systolic blood pressure (con), Diastolic blood pressure (con), Pulse (con), ECG abnormality (bin), Last mealtime (cat), Last meal type (cat) |
| Treatment | 1 | Average medication dosage (con) |
| CDSS | 9 | Calgary Depression Scale for Schizophrenia (con) |
| SWN-K | 20 | Subjective Well-being under Neuroleptic Treatment Scale (con) |
| MINI | 48 | Mini International Neuropsychiatric Interview |
| PANSS | 30 | Positive And Negative Symptom Scale (con) |
| PSP | 5 | Personal and Social Performance Scale (con) |
| CGI | 2 | Clinical Global Impression scale severity and improvement (con) |
| 10 weeks follow-up information used for determining the true remission status |  |  |
| PANSS | 8 | Positive And Negative Symptom Scale items: P1-3, N1, N4, N6, G5, G9 (con) |
| PSP | 5 | Personal and Social Performance Scale (con) |

con = continuous measure, bin = binary measure, cat = categorical measure.

#### Participants

Twelve psychiatrists and twelve residents in psychiatry, each with at least 1 year of clinical experience with severe mental illness, participated in the study. In the remainder of this paper, for the purpose of clarity and conciseness, all participants will be referred to as psychiatrists, unless stated otherwise. Written informed consent was obtained from all participants.

A pilot study showed that reaching a prognosis based on patient information was time-consuming for psychiatrists. To prevent participants from dropping out due to time constraints or the accuracy of predictions being affected by fatigue, we decided to divide the psychiatrists into three groups. Each participant group consisted of four psychiatrists and four residents in psychiatry. We aimed to distribute the level of experience in working with patients with psychotic disorders equally among the groups. The cases were also divided into three groups (group 1-3), with each group of psychiatrists being assigned one set of 22 cases.

#### Instruments

All participants completed a questionnaire (Supplement 2) in the Castor Electronic Data Capture online secure survey software program.^19^ All participants’ confidentiality agreements regarding the presented (pseudo-anonymized) patient data were obtained before participants commenced the real survey.

In the first part of the survey, general information about the participants (age, sex, country of birth, occupation (psychiatrist/resident in psychiatry) and years of experience working with patients with psychotic disorders) was collected.

For the second part of the survey, each participant group received baseline information on 22 (of the 66) randomly assigned patient cases from the OPTiMiSE dataset (Table 1, Supplement 1). For each case, participants predicted the chance of symptomatic and functional remission at 10 weeks on a scale of 0-100%. This first prediction is referred to as pre-MLM. Participants were also queried on the importance of specific patient information for their predictions.

Subsequently, the MLM prognosis and its level of certainty (uncertain, certain, definite) about that prognosis for the same cases,^18^ based on identical information, was presented to the participants, allowing them to adjust their predictions and provide reasoning. Whether adjusted or not, this second psychiatrists’ predictions are referred to as post-MLM.

The third part of the survey explored participants’ trust in artificial intelligence for prognosis prediction (measured on a 5-point scale), the extent to which they were inclined to consider the MLMs’ prediction (open-ended question), and whether they perceived any crucial information gaps in the patient data (open-ended question). Finally, psychiatrists were asked to estimate their predictive accuracy in this research on a scale between 0-100%.

### DATA-ANALYSIS

In a pre-analysis, we assessed whether the data of psychiatrists and residents in psychiatry could be pooled for further analysis. The means between the groups were compared with an independent two-sample t-test.

### 1. Who have better predictive performances; psychiatrists or MLM?

1a How accurate are the prognosis predictions of psychiatrists and MLM?

For each rater (participants and MLM), by comparing their predictions with the true outcomes, the following performance metrics were calculated: area under the receiver operating characteristic curve (AUC), accuracy, sensitivity, specificity, balanced accuracy and Brier score. Mean prognostic performances (across cases) of participants and MLM were compared using the non-parametric Mann-Whitney-U tests, because the participants’ data was not distributed normally for all prognostic metrics.

1b How comparable are the prognosis predictions among participants and between participants and the MLM?

Predictive agreements at group level (all participants + MLM) and pairwise interrater agreements were calculated with intraclass correlation coefficients (ICCs), based on a single-rating, absolute-agreement, 2-way random-effects model. In addition, the pairwise predictive agreement between participants and MLM was compared with the pairwise predictive agreement among participants by a Mann-Whitney-U test.

To visualize the relationships and distribution of accuracy of prognostic predictions of participants and MLM (for participants both pre-MLM and post-MLM), we calculated the Euclidean distances between the vectors of predictions for each group. Then we performed classical multidimensional scaling to construct rater representations in a 2-dimensional space, the coordinates of which were used for locating each rater in a scatter plot.

### 2. Can the MLM help participants to enhance the accuracy of their prognosis predictions?

2a Does the MLM help participants to enhance the accuracy of their predictions in general?

With a one-sample t-test we calculated whether the mean difference between accuracy of psychiatrists pre- and post-MLM significantly differed from 0.

2b Does the MLM help participants to enhance the accuracy of their predictions in cases with specific characteristics?

To effectively utilize the MLM, we aimed to identify cases where the psychiatrist’s prognosis predictions were poor, but the MLM performed well. All cases were grouped based on the number of correct predictions made by psychiatrists, creating nine difficulty groups ranging from 0 – 8 correct predictions. For each group, we calculated the percentage of correct predictions (accuracy) for both the psychiatrists and the MLM.

We then focused on the groups where psychiatrists made ≤ 3 correct predictions, defined as the ‘hard cases’.

For the ‘hard case’ groups, we calculated the expected number of correct predictions by both the psychiatrists and the MLM. The potential benefit of identifying cases in these groups was visualized.

Then, we investigated whether cases in these ‘hard case’ groups shared similar characteristics that could potentially be recognized by psychiatrists (or by computers). To do this, we created a t-distributed Stochastic Neighbor Embedding (t-SNE) plot to visualize the similarity of case characteristics. Data points representing ‘hard cases’ were highlighted with a distinct color to assess whether these cases shared more similar characteristics than others. More information about the characteristics and data-preparation used in this analysis is provided in Supplement 9.

### 3. How accurately do psychiatrists estimate their own predictive accuracy?

For both symptomatic and functional remission, the means of the psychiatrists’ estimated predictive accuracies were compared to their true predictive accuracies using a paired t-test. A Pearson correlation test was then performed to assess the strength and direction of the linear relationship between the estimated and true accuracies.

### 4. Do psychiatrists trust machine learning algorithms, and what influences their inclination to listen to the algorithm’s predictions?

The information participants provided on their level of trust in machine learning algorithms, and on what influenced their inclination to adhere to or deviate from the MLM predictions was summarized. For the questionnaire, see Supplement 2.

## FINDINGS

The mean age of the participants was 37.8 years (SD 9.1 years) and 50% were male. The mean number of years of working experience as a medical doctor in psychiatry was 15.1 (SD 8.5) year for psychiatrists and 4.0 (SD 1.4) years for residents.

### 0. Pre-analysis: Prognostic accuracy of psychiatrists and residents

A pooled variance t-test (Student’s t-test) showed no significant difference in mean accuracy between the resident and psychiatrist groups (symptomatic remission t(22) = -1.15, p = 0.26, and functional remission (t(22) = 0.18, p = 0.86).

### 1. Predictive performances of psychiatrists and MLM

Predictive performances of psychiatrists and the MLM are displayed in Table 2 and Supplement 3.

**Table 2.** Predictive performances of psychiatrists and the MLM.

|  | Psychiatrists |  | MLM |
| --- | --- | --- | --- |
|  | pre-MLM,<br>mean (SD) | post-MLM,<br>mean (SD) |  |
| Symptomatic remission |  |  |  |
| AUC | 0.58 (0.12) | 0.58 (0.10) | 0.59 |
| Accuracy | 0.52 (0.13) | 0.51 (0.13) | 0.50 |
| Sensitivity | 0.65 (0.22) | 0.71 (0.21) | 0.83 |
| Specificity | 0.44 (0.26) | 0.38 (0.27) | 0.24 |
| Balanced accuracy | 0.54 (0.11) | 0.55 (0.10) | 0.54 |
| Brier score | 0.28 (0.05) | 0.28 (0.05) | 0.28 |
| Functional remission |  |  |  |
| AUC |  |  |  |
| without group 1* | 0.76 (0.14) | 0.77 (0.13) | 0.77 |
| all participants | 0.70 (0.22) | 0.64 (0.26) | 0.67 |
| Accuracy |  |  |  |
| without group 1* | 0.75 (0.10) | 0.75 (0.11) | 0.80 |
| all participants | 0.72 (0.15) | 0.72 (0.15) | 0.79 |
| Sensitivity |  |  |  |
| without group 1* | 0.52 (0.27) | 0.55 (0.27) | 0.57 |
| all participants | 0.55 (0.36) | 0.41 (0.36) | 0.50 |
| Specificity |  |  |  |
| without group 1 | 0.79 (0.13) | 0.79 (0.13) | 0.84 |
| all participants | 0.75 (0.18) | 0.76 (0.18) | 0.83 |
| Balanced accuracy* |  |  |  |
| without group 1 | 0.65 (0.12) | 0.67 (0.13) | 0.70 |
| all participants | 0.65 (0.16) | 0.58 (0.17) | 0.66 |
| Brier score |  |  |  |
| without group 1 | 0.16 (0.05) | 0.16 (0.04) | 0.17 |
| all participants | 0.17 (0.05) | 0.17 (0.05) | 0.17 |

MLM = machine learning model, SD = standard deviation, AUC = area under the receiver operating characteristic curve
\*Because 'functional remission' occurred in only 1/22 of the randomly assigned cases in group 1, results of the sensitivity and therefore also AUC and balanced accuracy are not reliable/representative.

1a Comparison of predictive performances of psychiatrists versus MLM

The mean prognostic performances – AUC, accuracy, sensitivity, specificity, balanced accuracy and Brier score – of psychiatrists and MLM, for both symptomatic and functional remission, showed no significant differences (all p-values > 0.5).

1b Comparison of predictive agreement among psychiatrists and between psychiatrists and MLM Predictive agreement (ICC) of all raters (psychiatrist and MLM) at group level was in general poor with all group ICCs <0.5 for both symptomatic and functional remission (Table 3, Supplement 4 – Pairwise ICC matrices).^20^ For both symptomatic and functional remission the pairwise psychiatrist-MLM ICCs were significantly lower than psychiatrist-psychiatrist ICCs for group 3 (symptomatic remission p = 0.02, functional remission p = 0.001), but not for group 1 and 2.

**Table 3.**
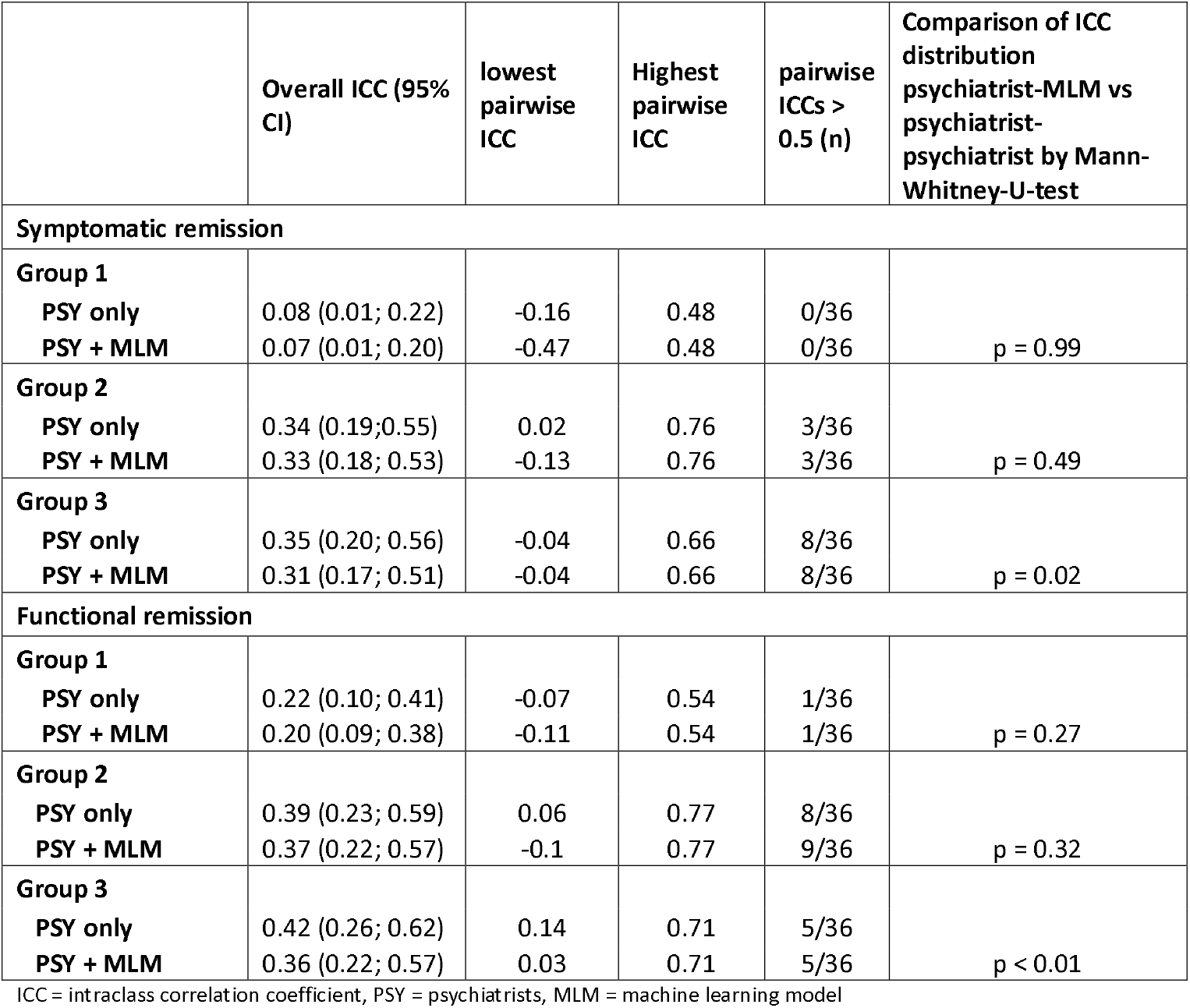
Predictive agreement by intraclass correlation coefficients (ICC)

Multidimensional Scaling (MDS) of the relationships and distributions of predictions on case-level for psychiatrists and MLM showed that the MLM’s predictions were largely similar to those of psychiatrists. In all plots, the MLM’s datapoint was positioned at the outer edge of the cloud of data points. However, in most plots there were psychiatrists who deviated from the general cloud as well. Psychiatrists’ predictions post-MLM were more similar (closer) to the MLM than those pre-MLM (Supplement 5).

### 2. Influence of the MLM on prognostic accuracy of psychiatrists

Predictive performances of psychiatrists pre-MLM and post-MLM are displayed in Table 2. About 25% of all predictions of psychiatrists were changed post-MLM. Some psychiatrists made no changes while others made changes in up to 77% of their predictions. The amount of percent change per prediction was highly variable as well, with the mean absolute amount of change per prediction ranging from 0 to 47%. Approximately the same amount of predictions was changed in the correct direction as in the incorrect direction post-MLM, both for symptomatic and functional remission (Supplement 6 and 7).

2a Influence of the MLM on mean prognostic accuracy of psychiatrists

The mean difference of accuracies of psychiatrists pre-MLM and post-MLM did not differ significantly from 0 for both symptomatic (t(23)= -0.84, p = 0.41) and functional remission (t(23)= -3.61e-16, p = 1) predictions.

2b Potential value of the MLM in cases that are difficult to predict for psychiatrists

The categorization of cases in groups based on how many psychiatrists predicted them correctly, showed that for functional remission many cases were easy to predict by the psychiatrists (nearly 50% of cases correct by ≥7 psychiatrists) while predicting symptomatic remission appeared more difficult for them (18% of cases correct by ≥7 psychiatrists).

For symptomatic remission, the ‘hard case’ groups (correct by ≤3 psychiatrists, 41% of cases) appeared to be more difficult to predict by the MLM as well (Figure 1). For functional remission, for the ‘hard cases’ groups (18% of cases) the accuracy of the MLM remained 50% or higher (Figure 1).

**Figure 1.**
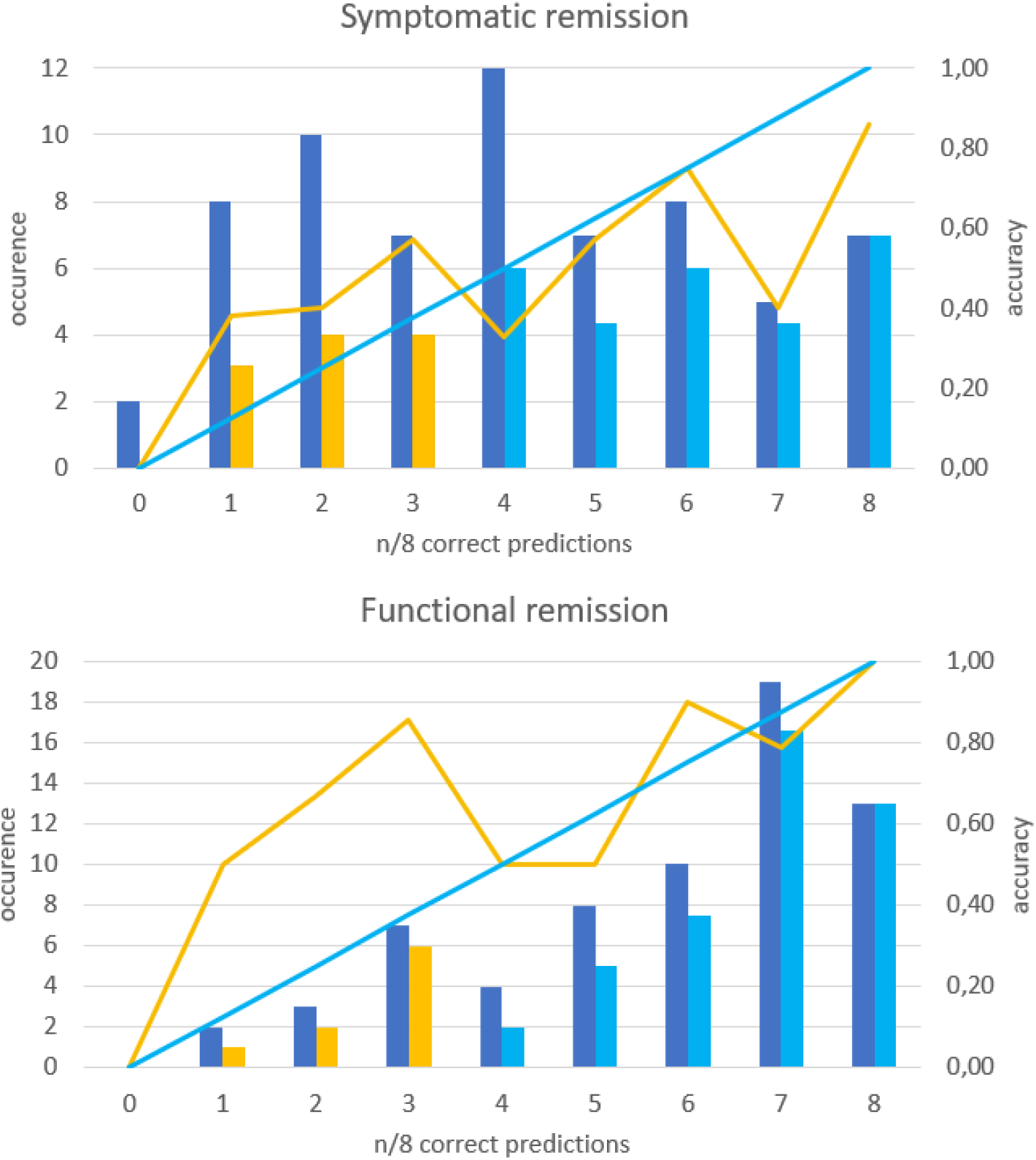
Potential benefit of using the MLM for the prediction of ‘hard cases’. The 66 cases are grouped based on the number of psychiatrists who correctly predicted them. The number of cases in each group is represented by the dark blue bars (‘occurrence’). Psychiatrist accuracy, calculated as the number of correct predictions divided by the total predictions for that group (group number/8), is shown by the blue line (‘psychiatrists’). The accuracy of the MLM for each group is represented by the orange line. For both symptomatic and functional remission, MLM accuracy (orange line) is higher than psychiatrist accuracy (blue line) for cases correctly predicted by < 4 psychiatrists, referred to as the ‘hard cases.’ ***Example:*** For the group of cases where 2 psychiatrists correctly predicted symptomatic remission, 10 cases (dark blue bar) were correctly predicted. Assuming psychiatrist performance reflects general practice, the expected number of correct predictions for this group is 2.5 (light blue bar), while the MLM correctly predicts 4 cases (orange bar). Thus, using the MLM instead of psychiatrists could result in 1.5 additional correct predictions, representing a 15% improvement.

For the ‘hard cases’, the MLM’s mean prognostic accuracy exceeded that of psychiatrists for both symptomatic and functional remission. The potential benefit of using the MLM for the prediction of these ‘hard cases’ is visualized in Figure 1.

The t-SNE plots showed that the case characteristics of the ‘hard cases’ were not distinct from the other cases (i.e., no outliers), making it impossible to identify the hard cases based on case characteristics (Supplement 8).

### 3. Psychiatrists’ estimation of their predictive accuracy

For symptomatic remission, psychiatrists overestimated their predictive accuracy (mean estimated accuracy = 0.62, mean true accuracy 0.52; t(19) = 2.79, p = 0.01). There was no significant correlation between the estimated and true accuracies (r = -0.19, p = 0.42). For functional remission, psychiatrists underestimated their predictive accuracy (mean estimated accuracy 0.55; mean true accuracy 0.72, t(19)= - 3.03, p < 0.01). Again, no significant correlation was observed (r= -0.23, p = 0.34).

### 4. The trust of psychiatrists in MLMs

Psychiatrists reported medium to high trust in the MLM, especially when it expressed higher certainty. When the MLM’s prediction differed substantially from their own, psychiatrists adjusted their predictions slightly in the direction suggested by the model. In response to open questions, three psychiatrists highlighted the MLM’s strength in processing large data sets and rendering impartial decisions. Six expressed reluctance to fully trust the model due to its lack of transparency. Seven noted the need for direct patient interaction for contextualization and interpretation of information. Quotes like, ‘You’re trying to turn a patient into statistics’ and ‘I treat patients, not questionnaires’, stressed the value of subjective elements like patient biography, motivation for treatment and the therapeutic alliance. Two psychiatrists viewed the MLM’s predictions as a second opinion for re-evaluating their decisions.

## DISCUSSION

### SUMMARY AND INTERPRETATION OF KEY FINDINGS

Accurate, personalized outcome prediction is essential for tailoring care to individual patients. This study explored whether machine learning models (MLMs) could improve prognosis prediction in first-episode psychosis using data from the OPTiMiSE study. We found that the mean predictive performance of the MLM was similar to that of psychiatrists for both symptomatic and functional remission. Interrater agreement among psychiatrists and between psychiatrists and the MLM was low but similar across groups.

Approximately 25% of psychiatrists’ predictions were altered after exposure to the MLM’s output. The interindividual variation in both the frequency and magnitude of these changes was substantial, with roughly equal proportions of corrections in the right and wrong direction. The MLM did not help to improve predictive accuracy of psychiatrists in general. However, Multidimensional Scaling (MDS) of the predictions revealed that although the MLM’s outputs were largely similar to those of psychiatrists, they often exhibited subtle deviations, as indicated by its position at the outer edge of the cloud. This slightly different predictive approach may help explain why the MLM occasionally outperformed psychiatrists in hard cases. A critical issue was psychiatrists’ difficulty in recognizing when to rely on the model and we could not identify a clear pattern to identify ‘hard cases’ based on their characteristics.

Psychiatrists tended to overestimate their accuracy for predicting symptomatic remission, while underestimating accuracy for functional remission. These biases may reflect the routine assessment of symptom severity and less frequent evaluation of functional recovery in clinical practice. This discrepancy in self-perceptions likely influenced how psychiatrists engaged with the MLM’s outputs, shaping their reliance on and interpretation of the model’s predictions.

While psychiatrists expressed moderate to high trust in MLMs for prognosis prediction, they voiced concerns about the lack of transparency regarding how the model made predictions. For MLMs to be useful, they must be interpretable.^21^ Explainable artificial intelligence (XAI) techniques, such as feature importance measures,^22 23^ can highlight the variables most crucial for predictions, could improve data-driven explanations and aid in shared-decision making with patients.^24-26^

Surprisingly, we found no difference in accuracy between psychiatrists and psychiatry residents, which contrasts with previous studies suggesting that clinical experience improves decision-making accuracy.^27^ One explanation for this discrepancy could be the absence of direct patient interaction in this study, which prevented participants from forming a ‘clinical impression’. The clinical impression might get better with increasing experience, improving predictive accuracy in real-world setting.

### LIMITATIONS

First, the OPTiMiSE dataset lacked key clinical predictors such as trauma history^28^, motivation to and adherence to treatment^29-31^ and pre-morbid functioning,^32^ which may have affected prediction accuracy. Second, psychiatrists in this study relied solely on case data, lacking direct patient interaction, which could have influenced their predictions. Third, the small sample size and low interrater agreement may have limited the detection of small effects. Due to the random assignment, group 1 contained only one patient that achieved functional remission, affecting the reliability of calculations for this group.

Fourth, the 10-week follow-up may have been too short to achieve functional remission, which typically takes more time.^33^ Finally, outcome measures (PANSS and PSP scores) were selected and rated by researchers and clinicians, not patients.

### RECOMMENDATIONS FOR FUTURE RESEARCH

Incorporating a broader range of prognostic factors may improve the predictive accuracy of both psychiatrists and MLMs. Machine learning techniques from fields such as weather forecasting,^34^ which successfully intergrate complex data, may offer strategies to improve psychiatric predictions. Future MLMs incorporating multimodal data such as neuroimaging, EEG, genetics and wearable data may enhance accuracy. Longitudinal data could also refine predictions.^18^ However, increasing the data collection burden might limit the model’s feasibility for routine clinical use.

Future research should also explore how direct clinical contact influences prognostic accuracy by comparing predictions based on case data with those made after in-person evaluations. If ‘clinical impression’ is found to predict outcomes, methods for integrating this into MLMs should be developed. One approach is to provide MLMs with descriptive diagnoses, which offer a more comprehensive and holistic understanding of the patient’s condition, aligning more closely with clinicians’ impressions than standard disease classifications. In such a descriptive diagnosis, it would be important to also incorporate the perspective of patients and their informal caregivers, because previous research showed that the perspectives of patients, informal caregivers and healthcare professionals on barriers and facilitators of recovery can diverge and possibly complement each other.

Larger sample sizes and more balanced case distributions will also be important for detecting subtle effects, particularly in outcomes like functional remission. Extending the follow-up period beyond 10 weeks is crucial to capture long-term recovery, especially for functional outcomes. For personalized treatment, future research should prioritize patient-relevant outcomes measures. ^35 36^ Finally, comparing the decision-making processes of psychiatrists and MLMs could shed light on how they can complement each other.

### CLINICAL IMPLICATIONS

Despite the comparable predictive performance, replacing psychiatrists with MLMs in prognosis prediction tasks is unlikely, due to the clinical responsibility and liability associated with medical decisions. A more realistic approach would involve using MLM predictions as a second opinion, as we did in this study. In this scenario, psychiatrists would retain full autonomy over their final decision, integrating the MLM output as one piece of supplementary information. Since we do not know in which cases psychiatrists should rely on the MLM, a potential strategy could be to consistently average the predictions of the psychiatrist and the MLM. This approach leverages the strengths of multiple sources to mitigate individual biases and errors.^37-39^ In the current study, this would have resulted in a mean post-MLM accuracy of 0.54 for symptomatic remission (actual values pre-MLM 0.52, post-MLM 0.51, MLM 0.50) and 0.79 for functional remission (actual values pre-MLM 0.72, post-MLM 0.72, MLM 0.79).

For MLMs to add value in clinical practice, several improvements are necessary. First, increasing the accuracy of the MLM would enhance its utility in refining psychiatrists’ predictions. Second, identifying when psychiatrists should rely on the MLM is critical. This could be achieved by improving the model’s ability to estimate the certainty of its predictions or by identifying cases that are hard to predict for psychiatrists. Additionally, explaining model decisions would allow psychiatrists to make more informed judgments about when to trust the model’s output.

## Acknowledgements

ChatGPT (GPT-4, October 2024 version) was used to review and improve the grammar, language, and flow of the manuscript.

## Ethics statement

The authors assert that all procedures contributing to this work comply with the ethical standards of the relevant national and institutional committees on human experimentation and with the Helsinki Declaration of 1975, as revised in 2013. All procedures involving human subjects were approved by the Ethics Committee of the University Medical Centre Utrecht (protocol number 20/053).

## Conflicts of Interest

None declared

## Data availability statement

The data that support the findings of this study and the analytic code are available from the corresponding author, [VvD], upon reasonable request.

## Funding

This work was supported by ZonMw (project ID 63631 0011) and by a grant from the working group AI for Health of the Alliance TU/eWUR-UU-UMCU for the project.

## Authors contribution

VD, SK, CF, WS, WC and HS contributed to the conception and design of the study. AA, AB, CB, DC, ED, LDij, AD, LDo, JE, FG, MH, FH, CK, LK, ML, MK, BM, DR, ER, RS, MS, JS, MV, JV participated as participants (residents/ psychiatrists) in the study. VD, SK, WS, WC and HS drafted the manuscript. RK and IW-vR provided the data and organized the database. VD, HS and SK carried out the modeling and statistical analysis. All authors critically reviewed the article and approved the submitted version.

## Notes

### Competing Interest Statement

The authors have declared no competing interest.

### Author Declarations

All procedures involving human subjects were approved by the Ethics Committee of the University Medical Centre Utrecht (protocol number 20/053).

