## Supplement for "Prognostic predictions in psychosis: exploring the complementary role of machine learning models"

**Supplemental materials**

**Content**

| **Supplemental material** | **Page(s)** |
| --- | --- |
| Supplement 1 - Characteristics of the included 66 patients from the OPTiMiSE trial | 2 |
| Supplement 2 Example of Castor survey ‘Psychiatrists versus machine in psychosis prognosis prediction’ | 3-6 |
| Supplement 3 - Predictive performances of psychiatrists and MLM | 7-10 |
| Supplement 4 - Interrater agreement by intraclass correlation coefficients (ICCs) | 11-12 |
| Supplement 5 - Relationships and distributions of predictions by multidimensional scaling plots | 13-14 |
| Supplement 6 - Changes in predictions by psychiatrists post-MLM | 15 |
| Supplement 7 - Visualisation of accuracy of predictions pre- and post-MLM | 16-17 |
| Supplement 8 - Relative simillarity between cases based on patient characteristics | 18-19 |

**Supplement 1 - Characteristics of the included 66 patients from the OPTiMiSE trial**

| ***Baseline*** |  |
| --- | --- |
| Sex, male | 79% |
| Mean age, years (SD) | 25.3 (0.8) |
| DSM-classification |  |
| - Schizophrenia | 67% |
| - schizophreniform disorder | 32% |
| - schizo-affective disorder | 2% |
| Inpatient status | 55% |
| Mean duration of currect psychotic episode, months (SD) | 2.5 (0.5) |
| (Volunteer)work or school | 32% |
| Psychiatric comorbidities | 30% |
| Symptomatic remission according to RSWG-criteria* | 0% |
| Functional remission defined as PSP >70 | 12% |
| ***Outcome at week 10*** |  |
| Symptomatic remission according to RSWG-criteria* | 44% |
| Functional remission defined as PSP >70 | 12% |

*only the symptom severity component of the RSWG criteria was used, not the time-component.

**Supplement 2 - Example of Castor survey ‘Psychiatrists versus machine in psychosis prognosis prediction’**

Introduction

The following survey contains sensitive information about patients. By clicking the 'Next' button, the participant agrees not to disclose any information about the questionnaire to third parties and to close all forms containing sensitive information after completing them.

General respondent information

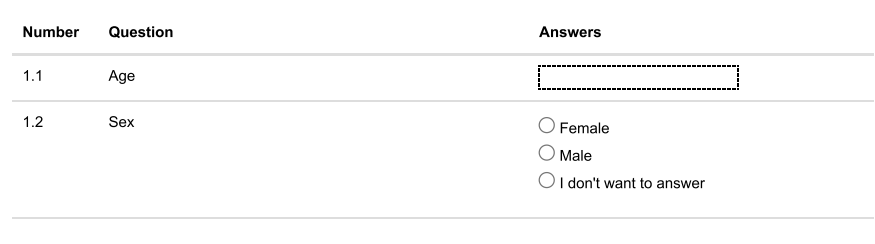

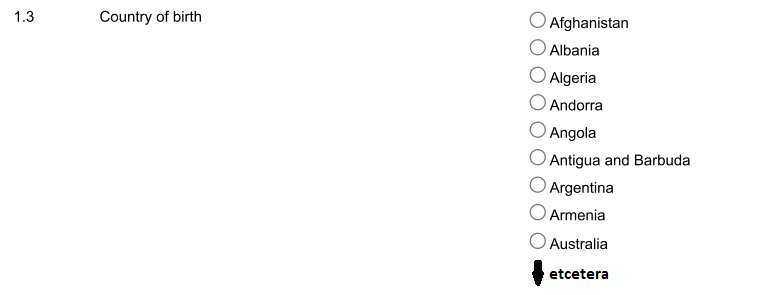

Case 1 - Patient information

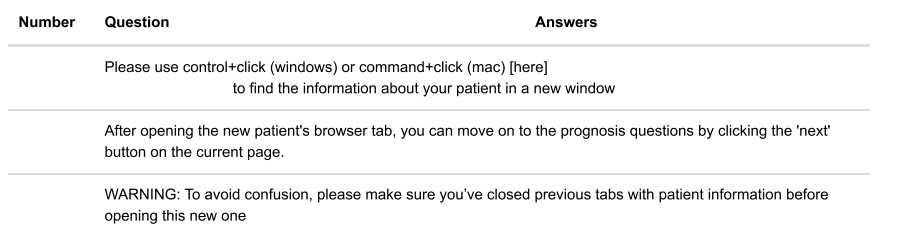

Case 1 – Prediction pre-MLM

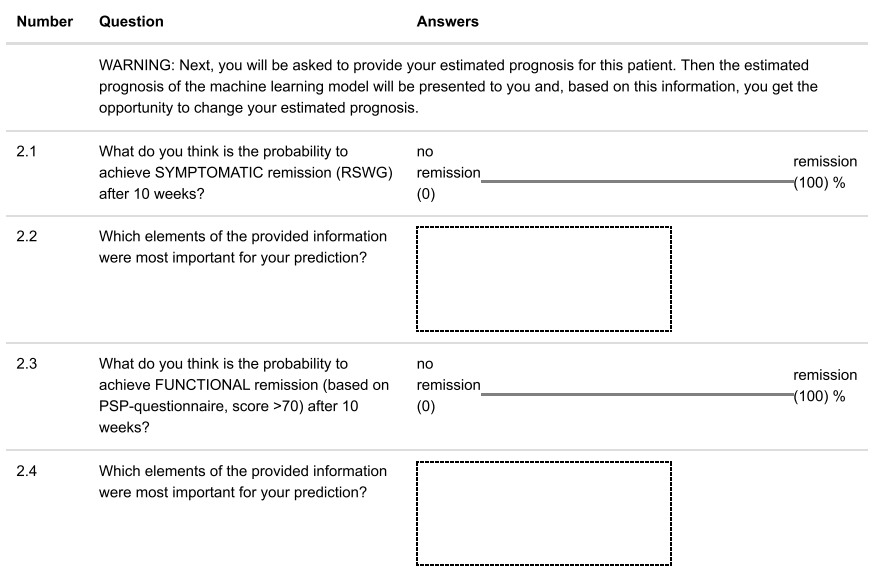

Case 1 – Prediction post-MLM

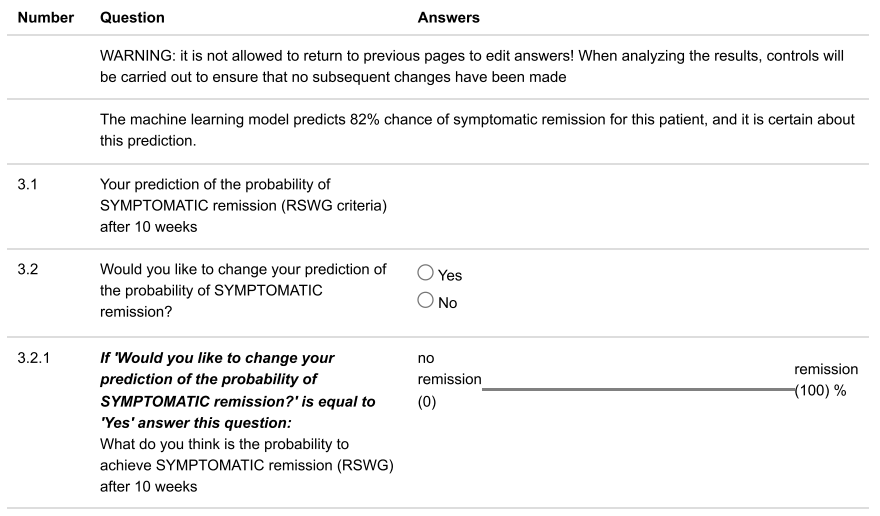

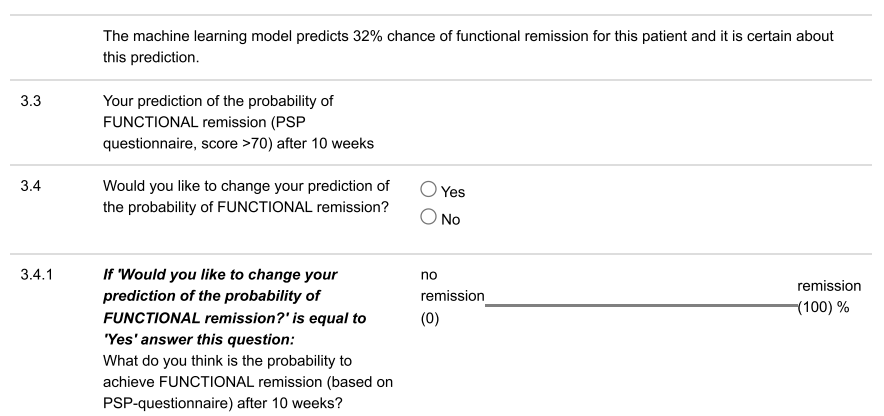

Estimated predictive accuracy

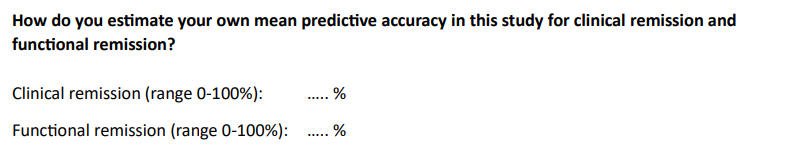

Final information – Artificial Intelligence

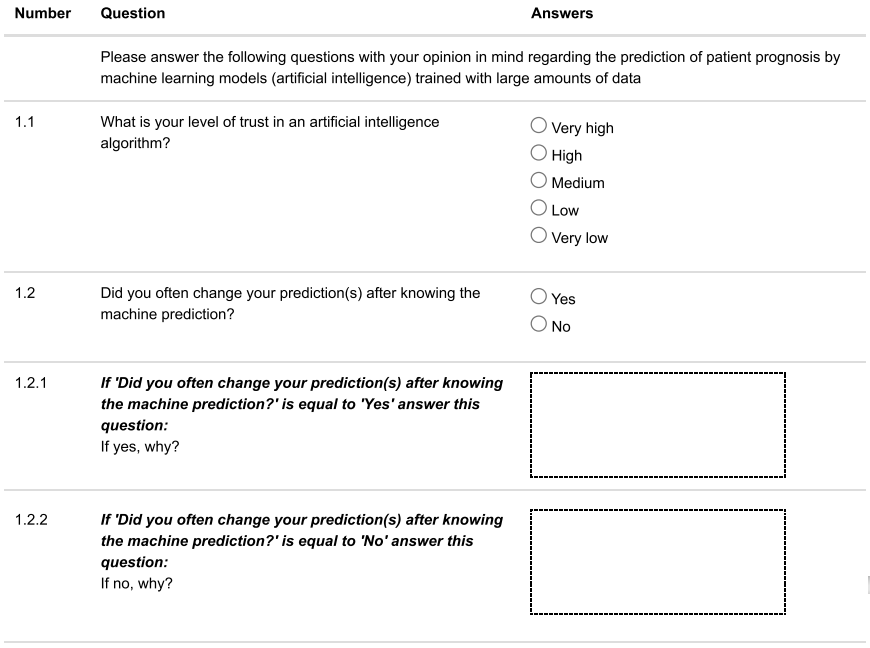

Final information – Ecological value

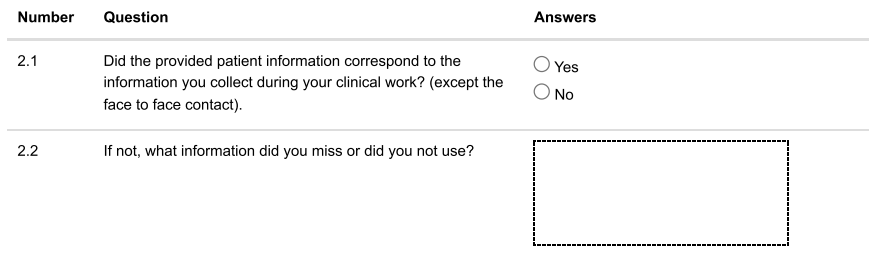

Final information – Fatigue

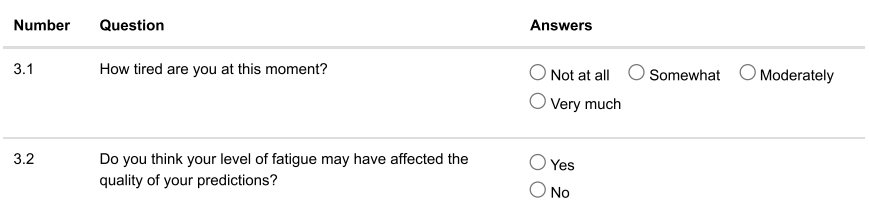

Final information – End of questionnaire

Thank you for participating in our survey

**Supplement 3 - Predictive performances of psychiatrists and MLM**

Participants are labeled by group number (G1-4) and psychiatrist (P) or resident (R) status

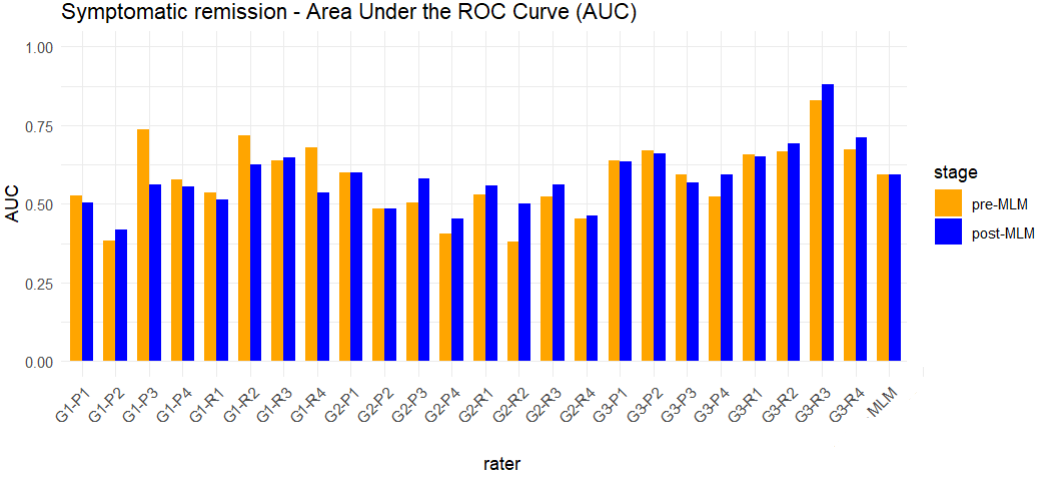

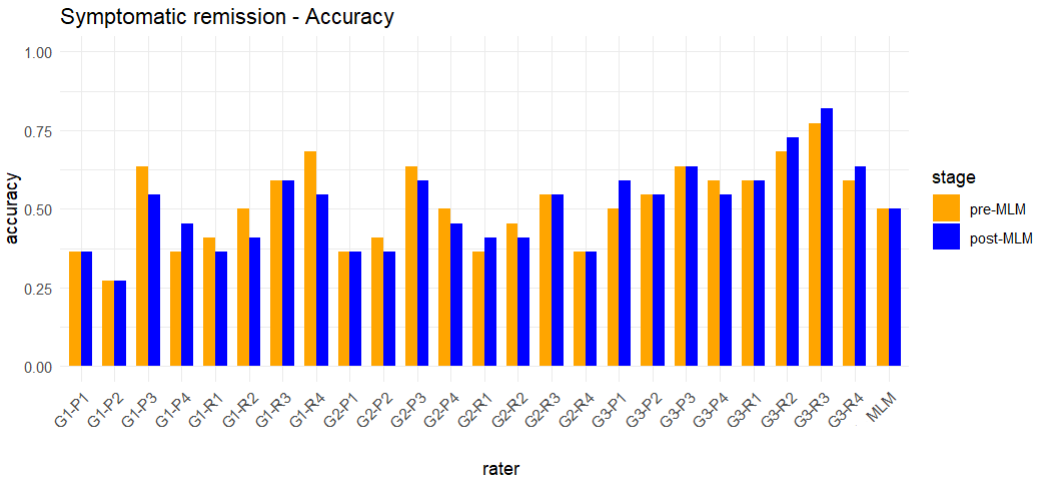

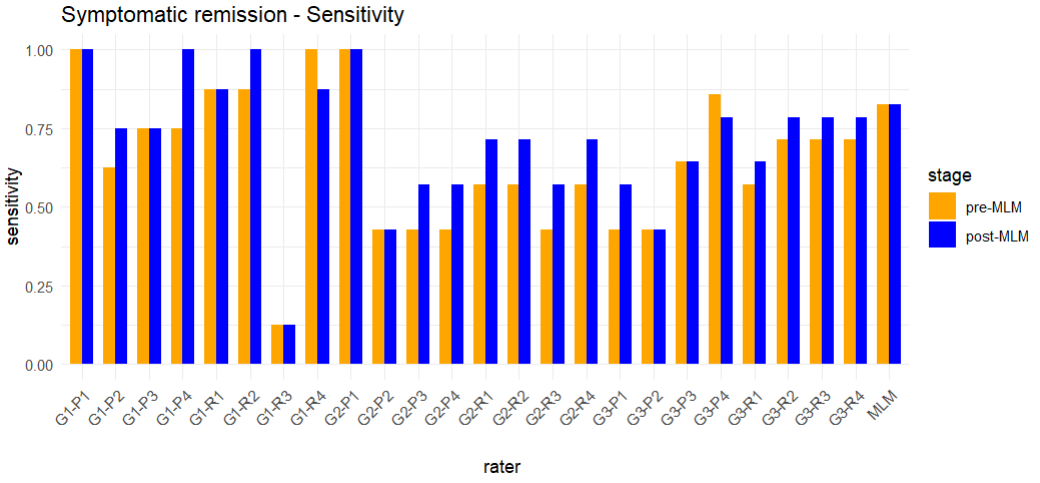

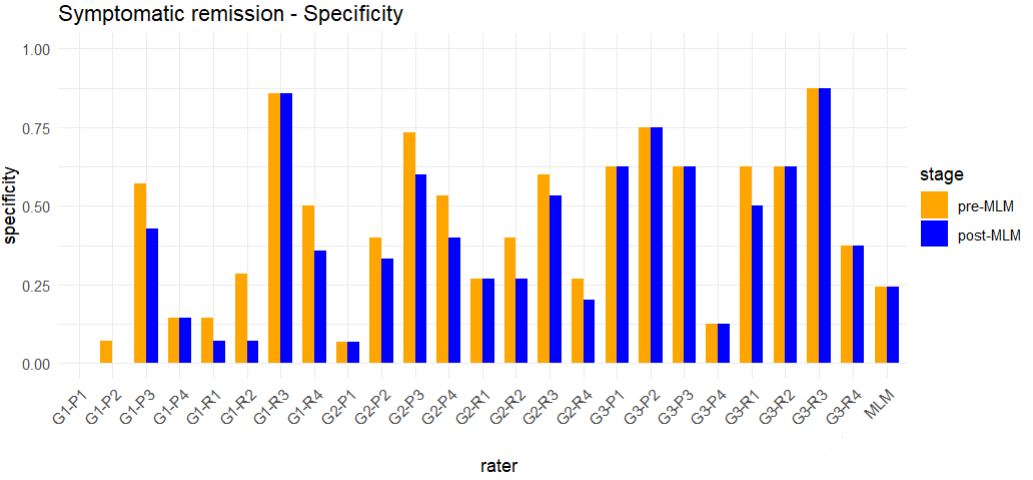

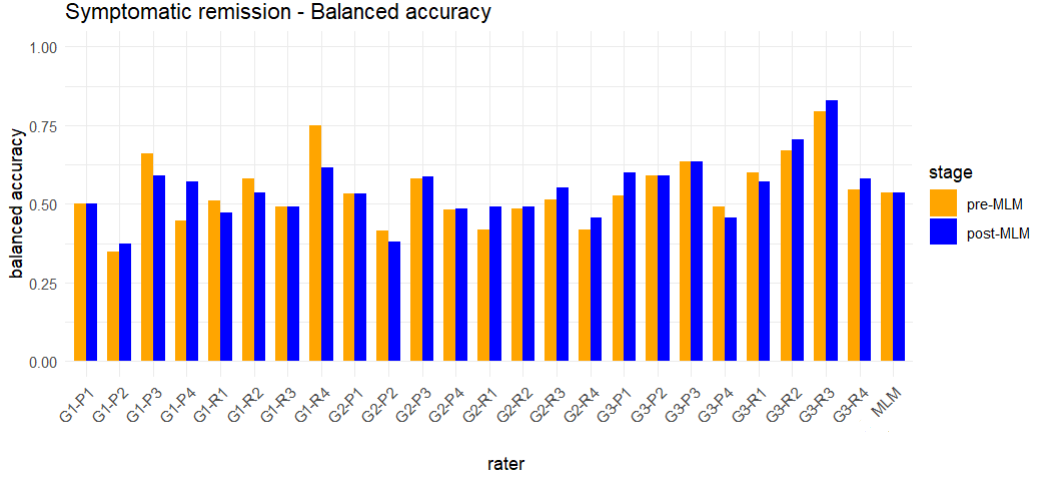

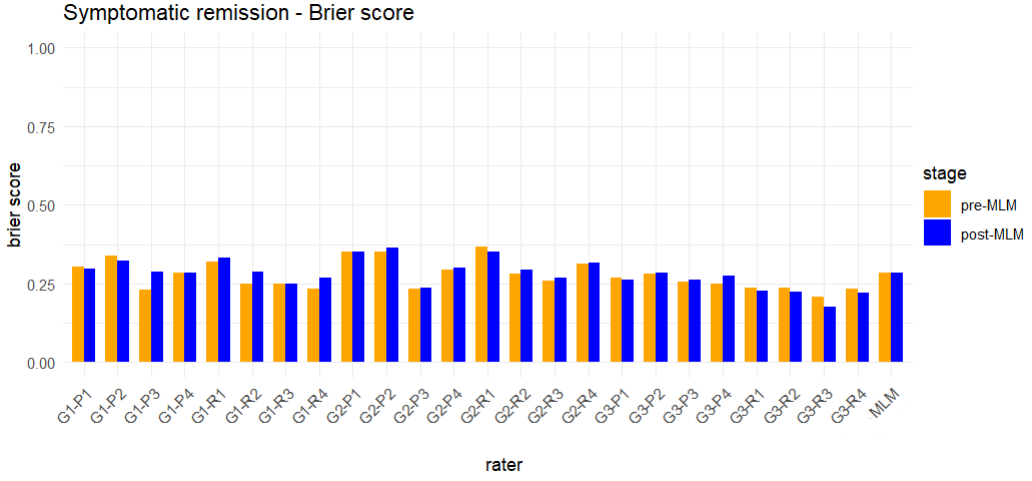

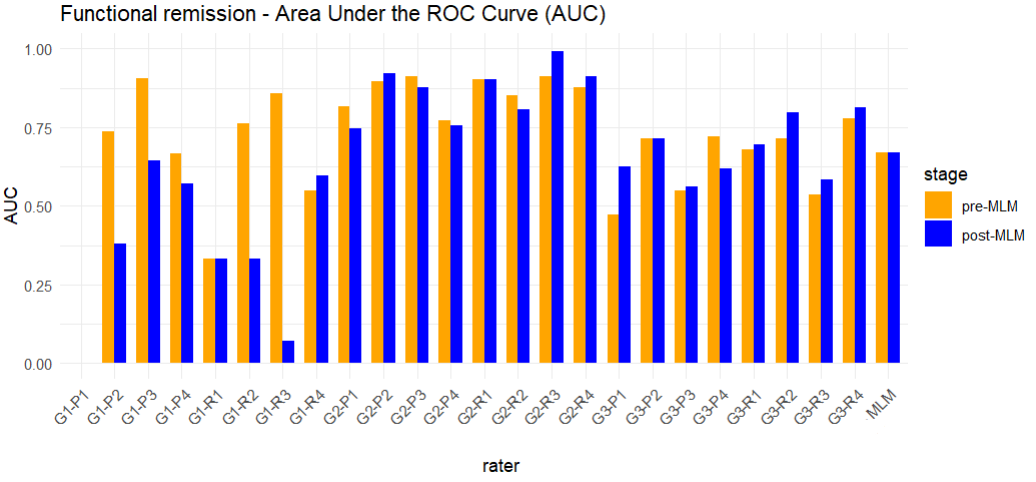

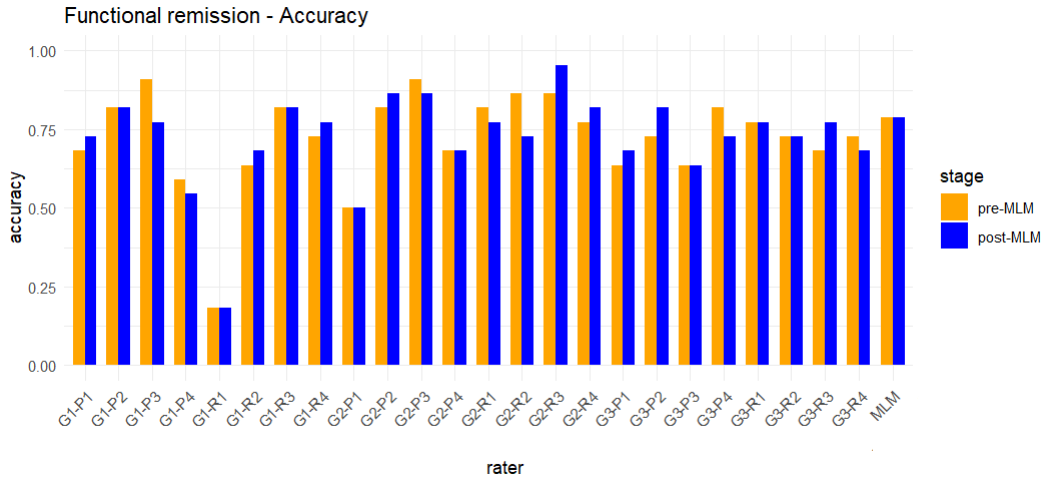

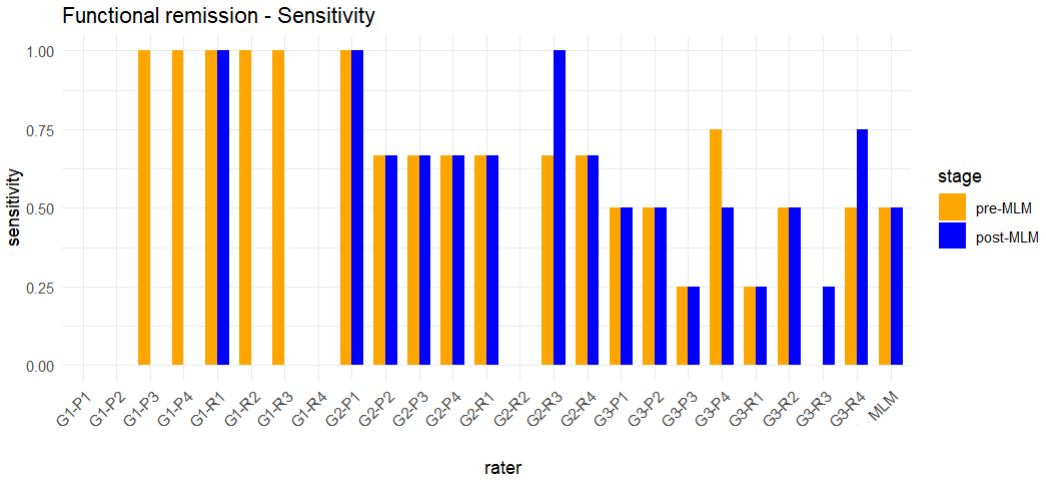

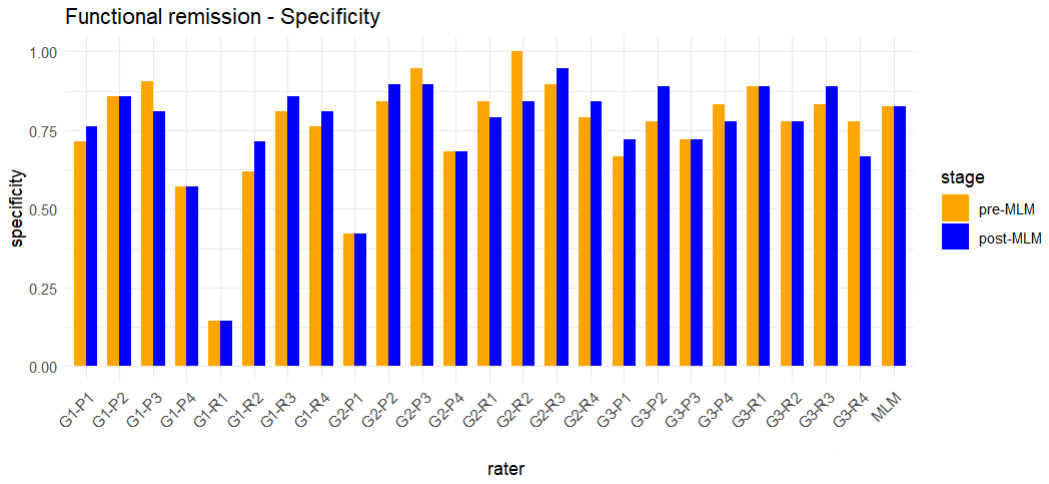

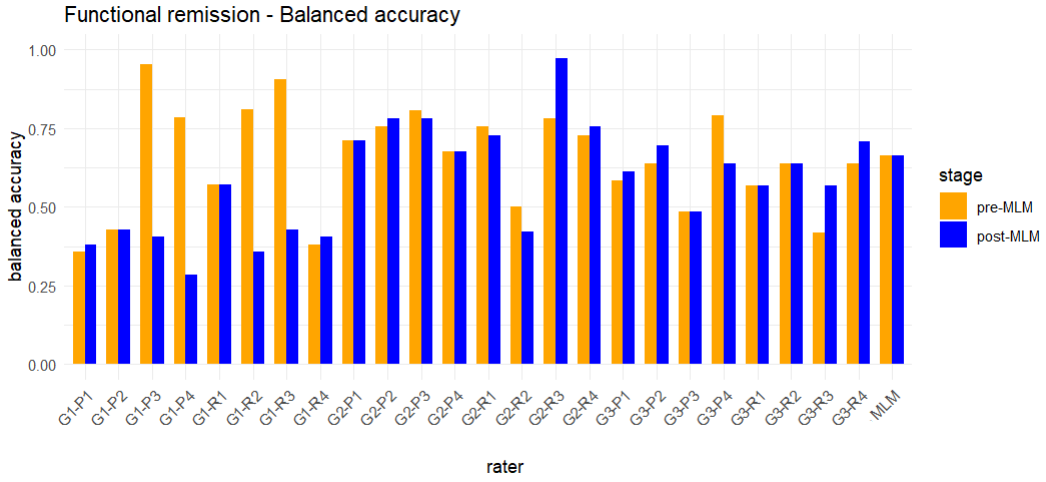

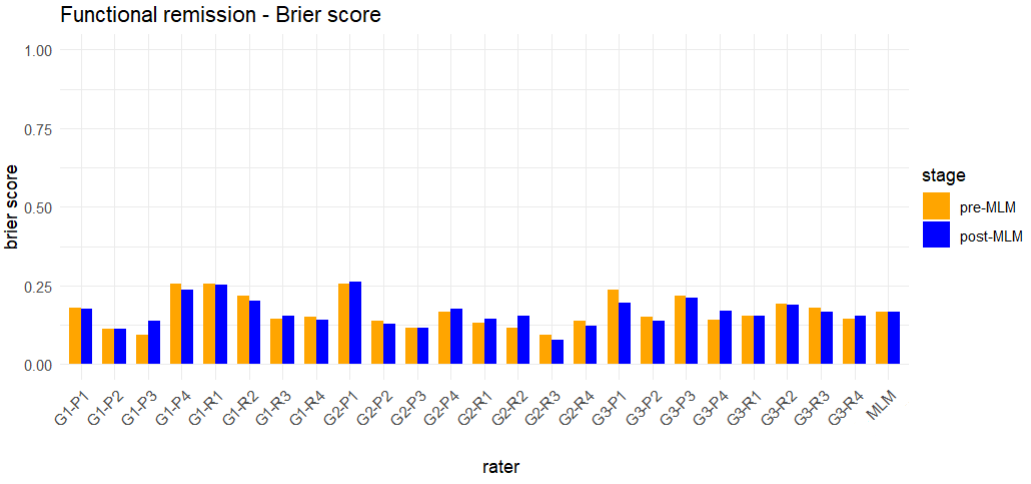

**Supplement 4 - Interrater agreement by intraclass correlation coefficients (ICCs)**

Participants are labeled by group number and psychiatrist or resident status

**Symptomatic remission**

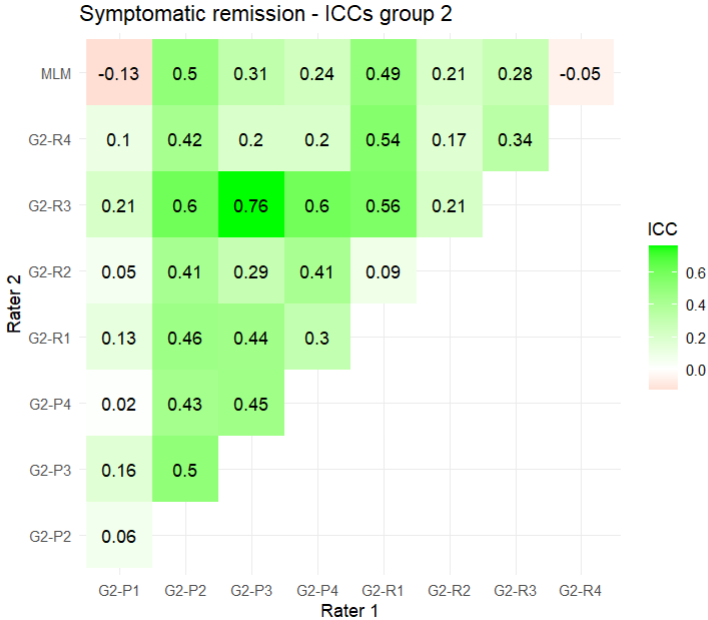

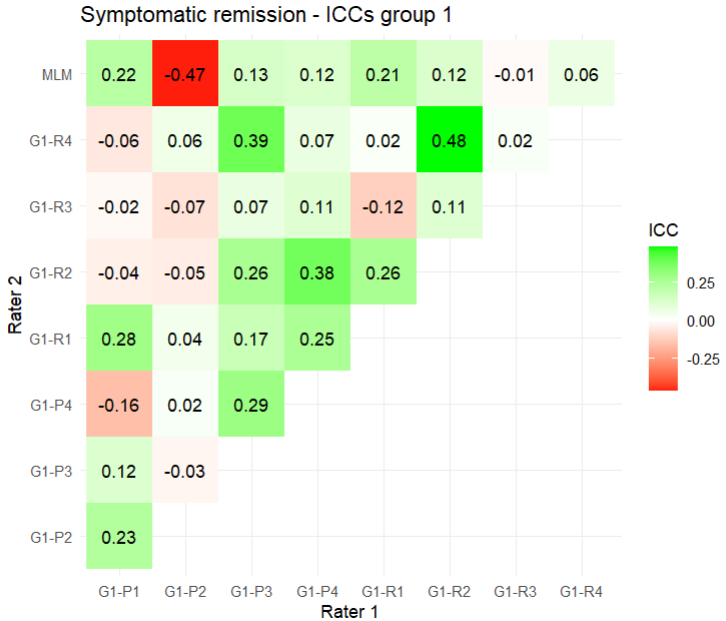

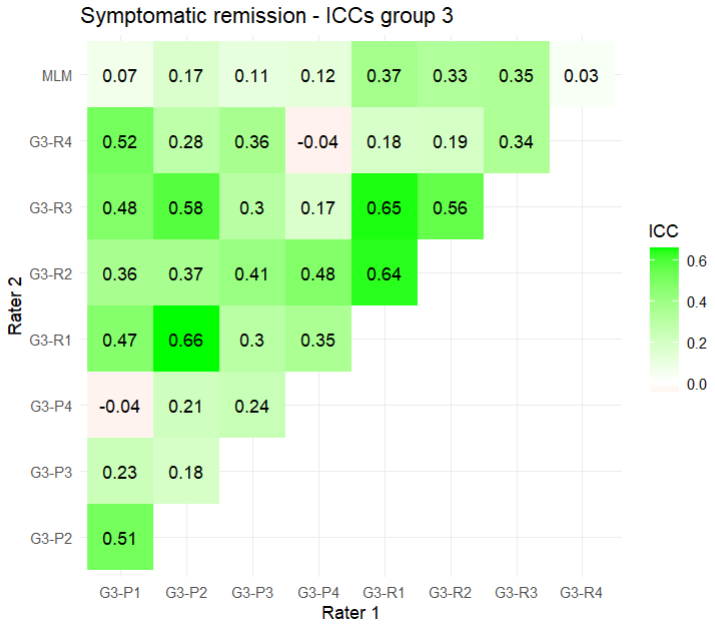

**Functional remission**

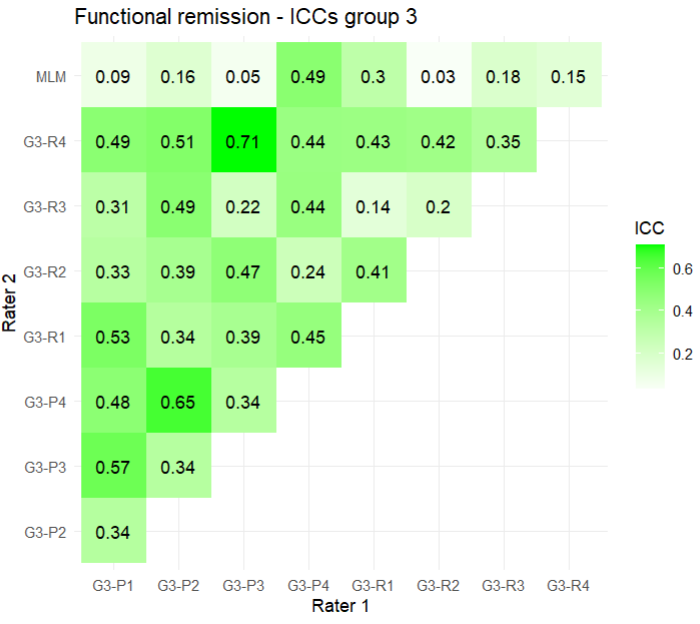

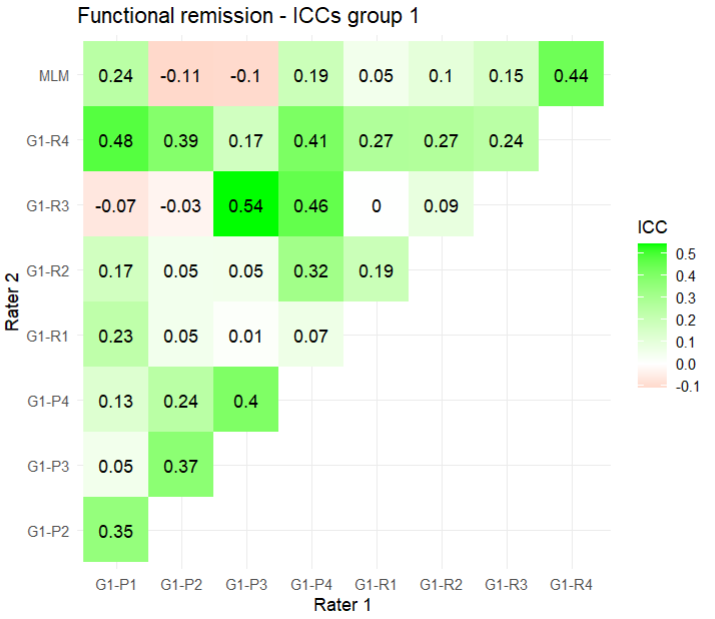

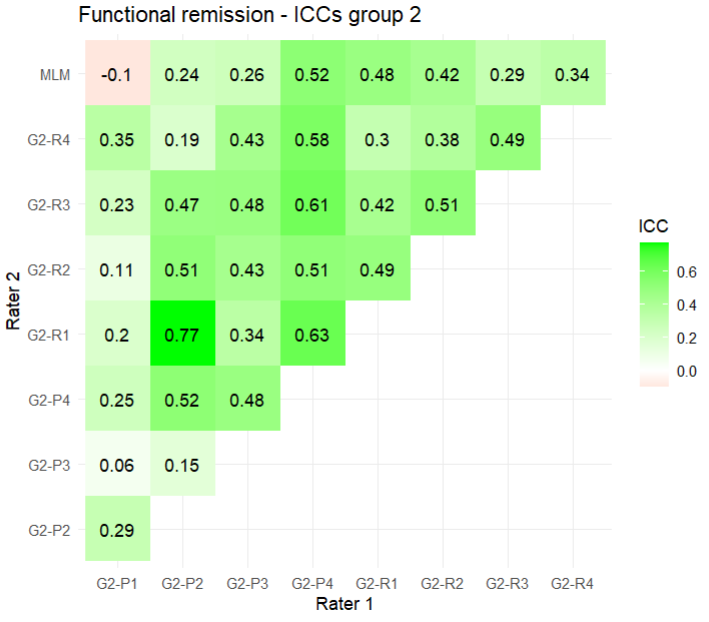

**Supplement 5** **- Relationships and distributions of predictions by multidimensional scaling plots**

PSY pre-MLM	PSY post-MLM	MLM

Each psychiatrist is represented by a different color.

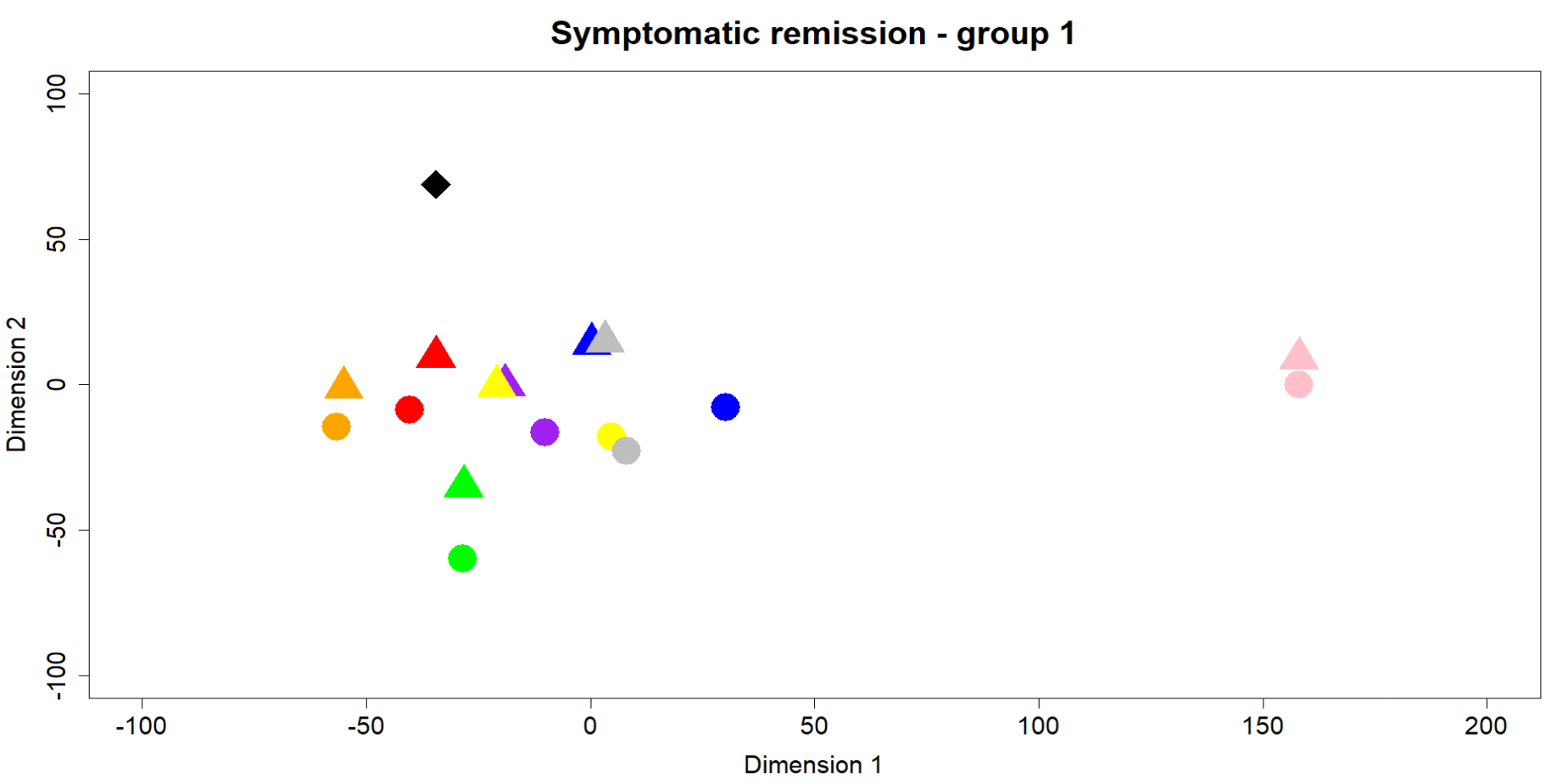

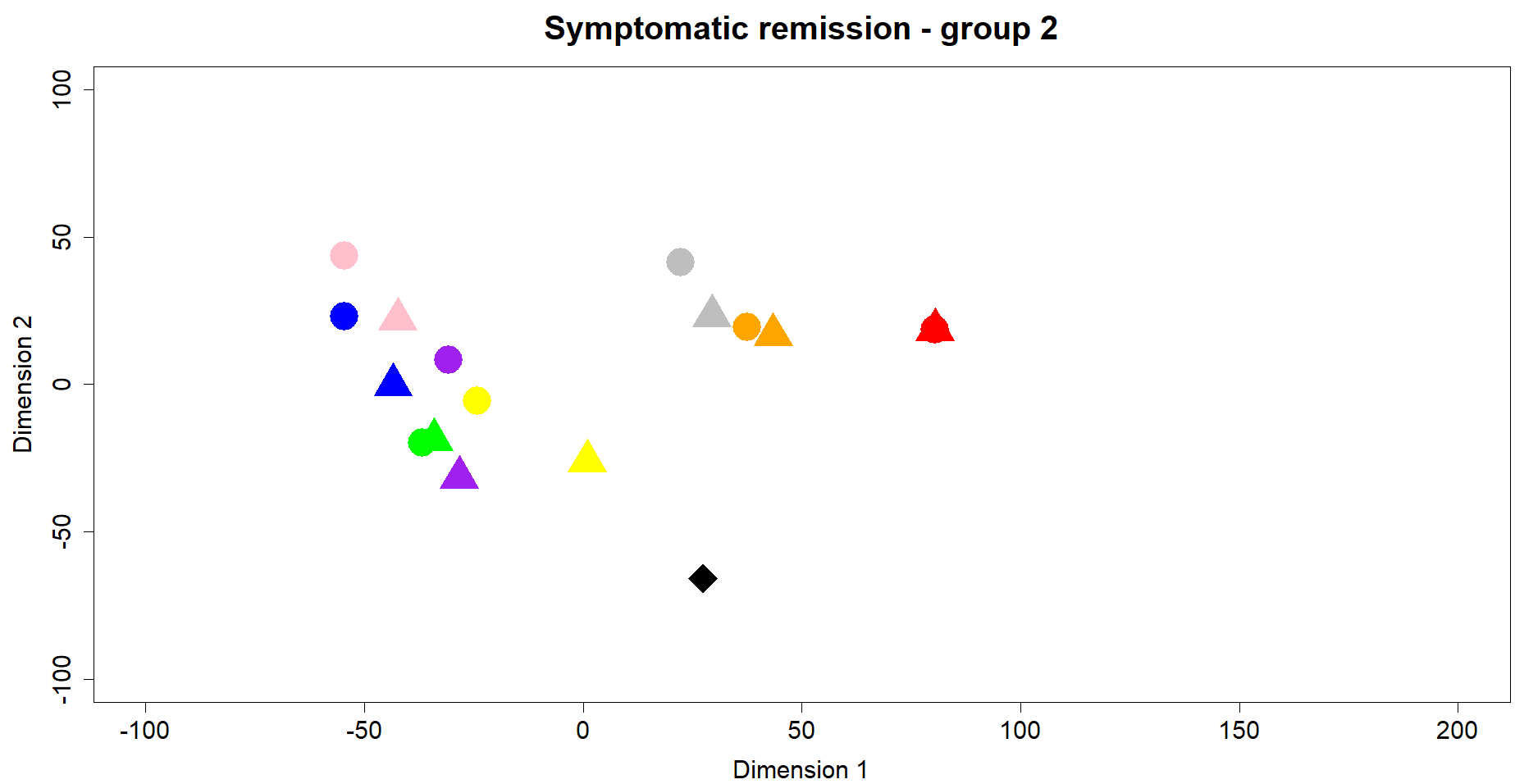

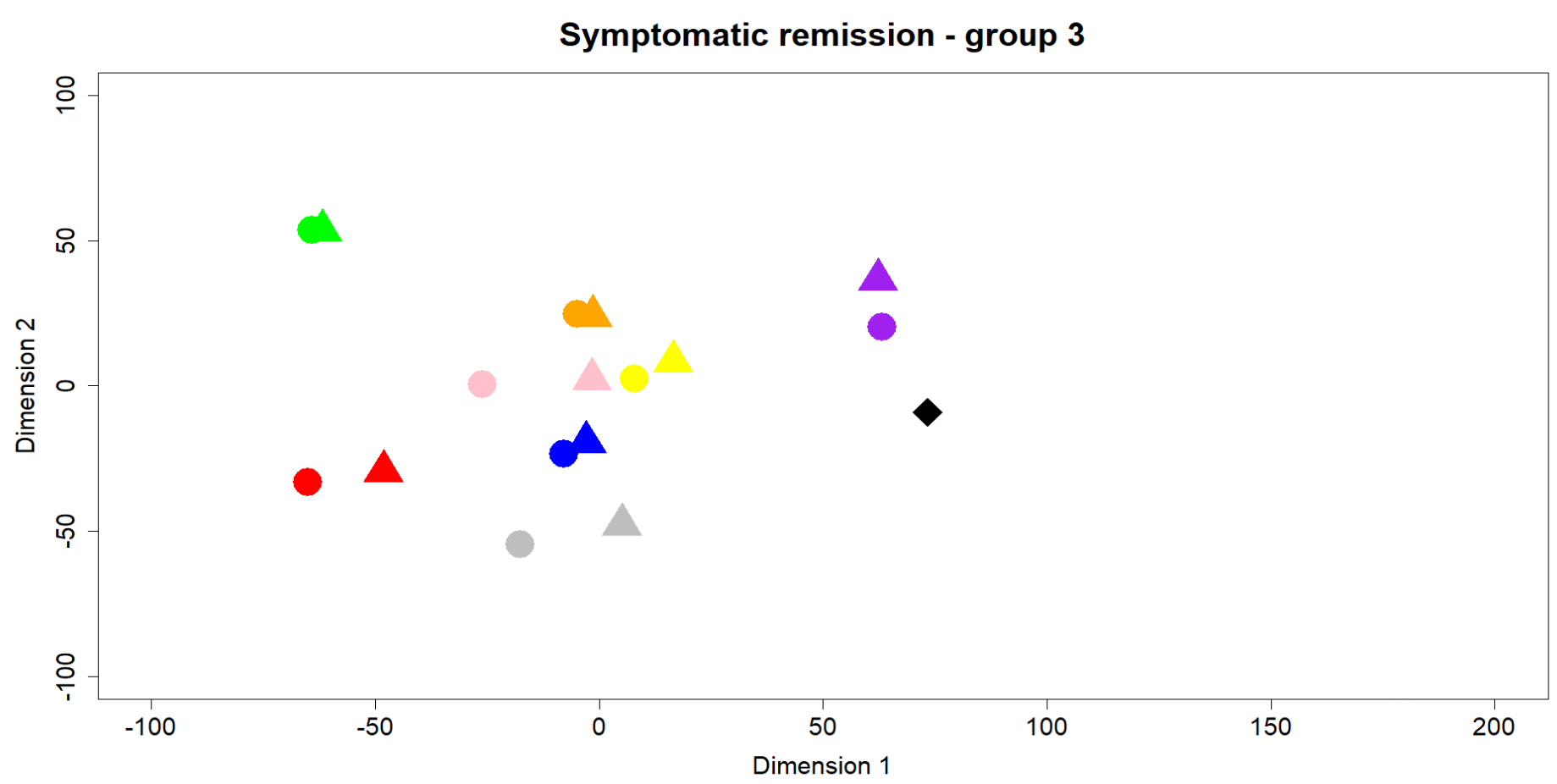

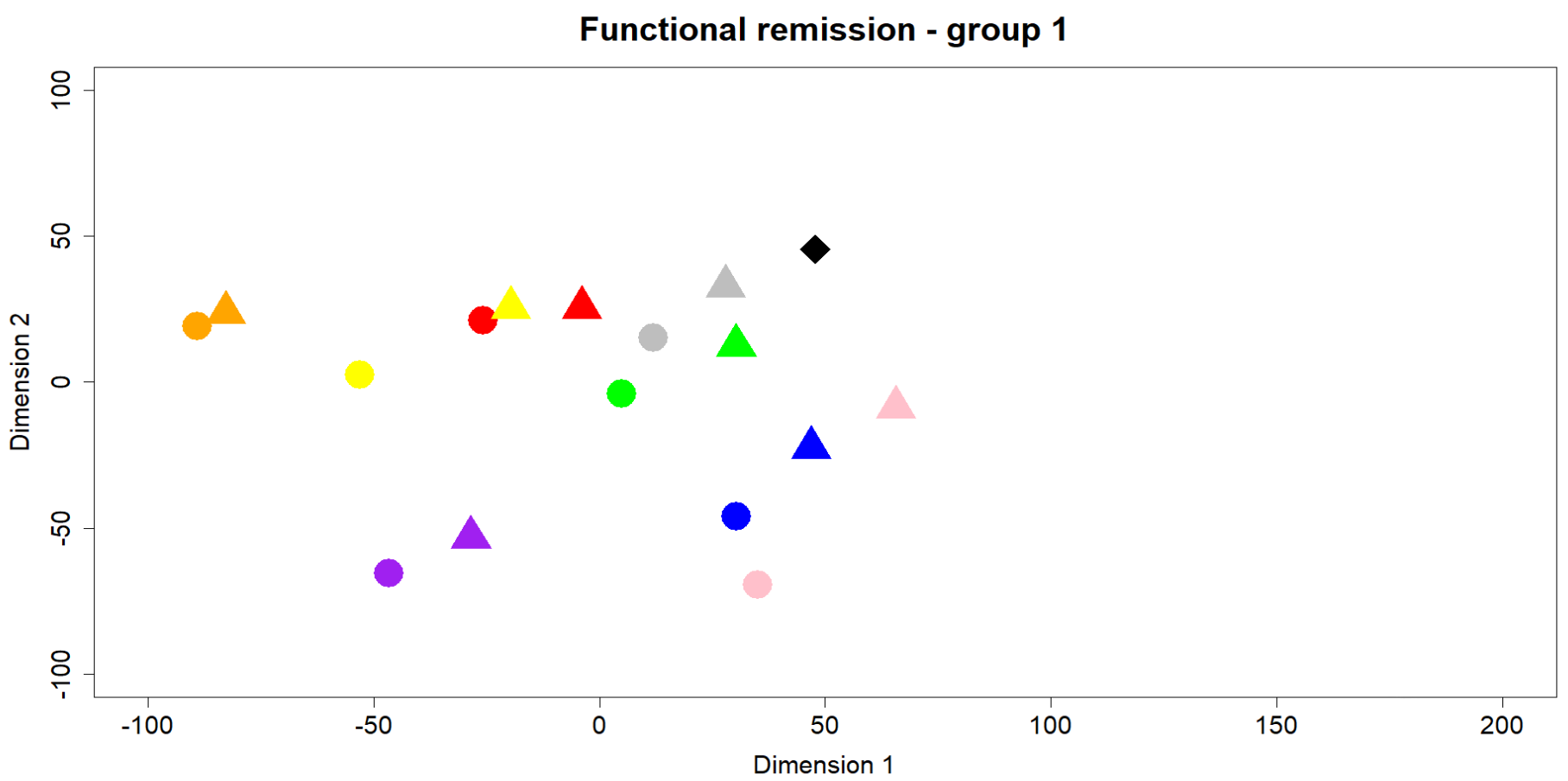

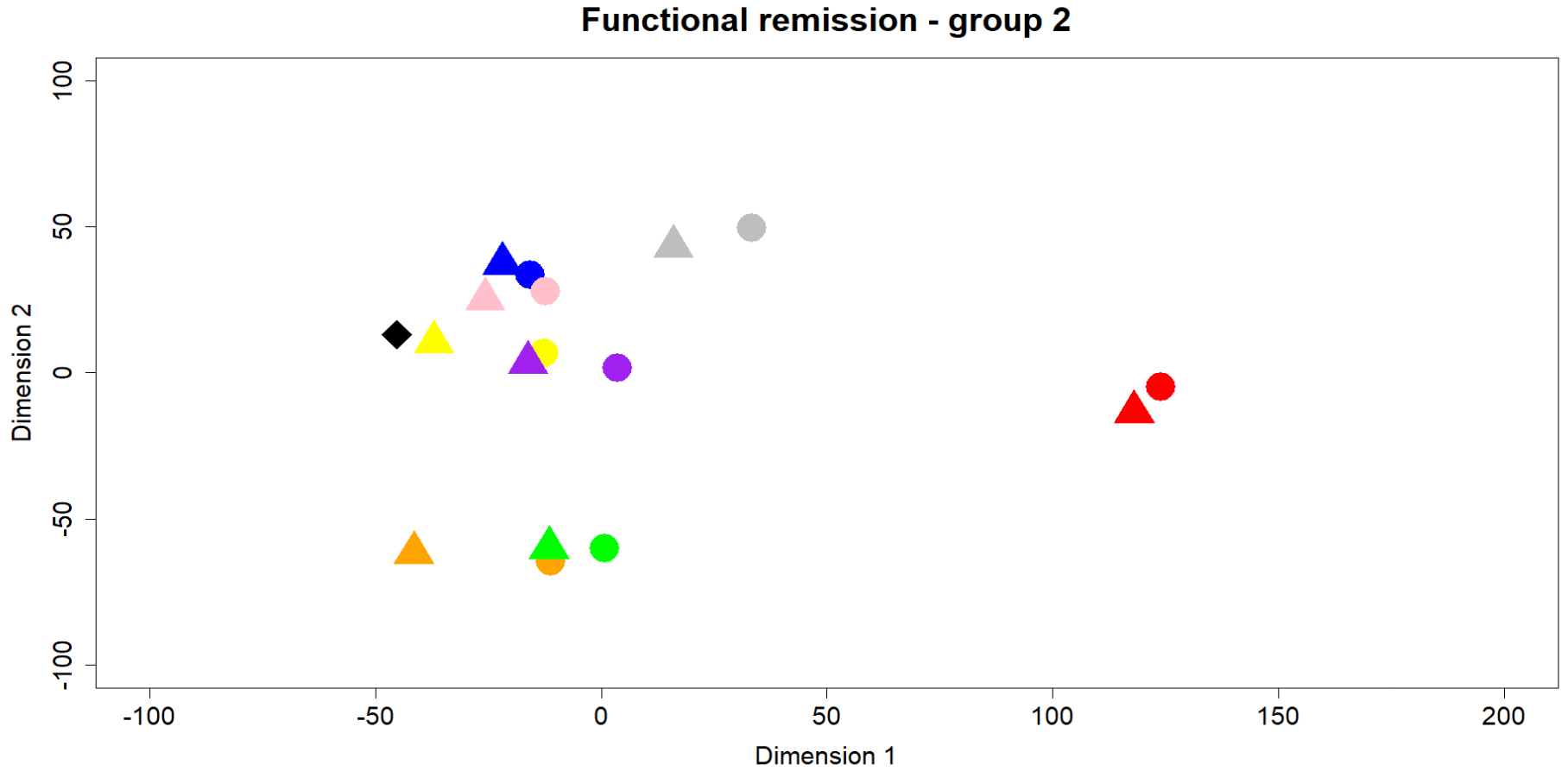

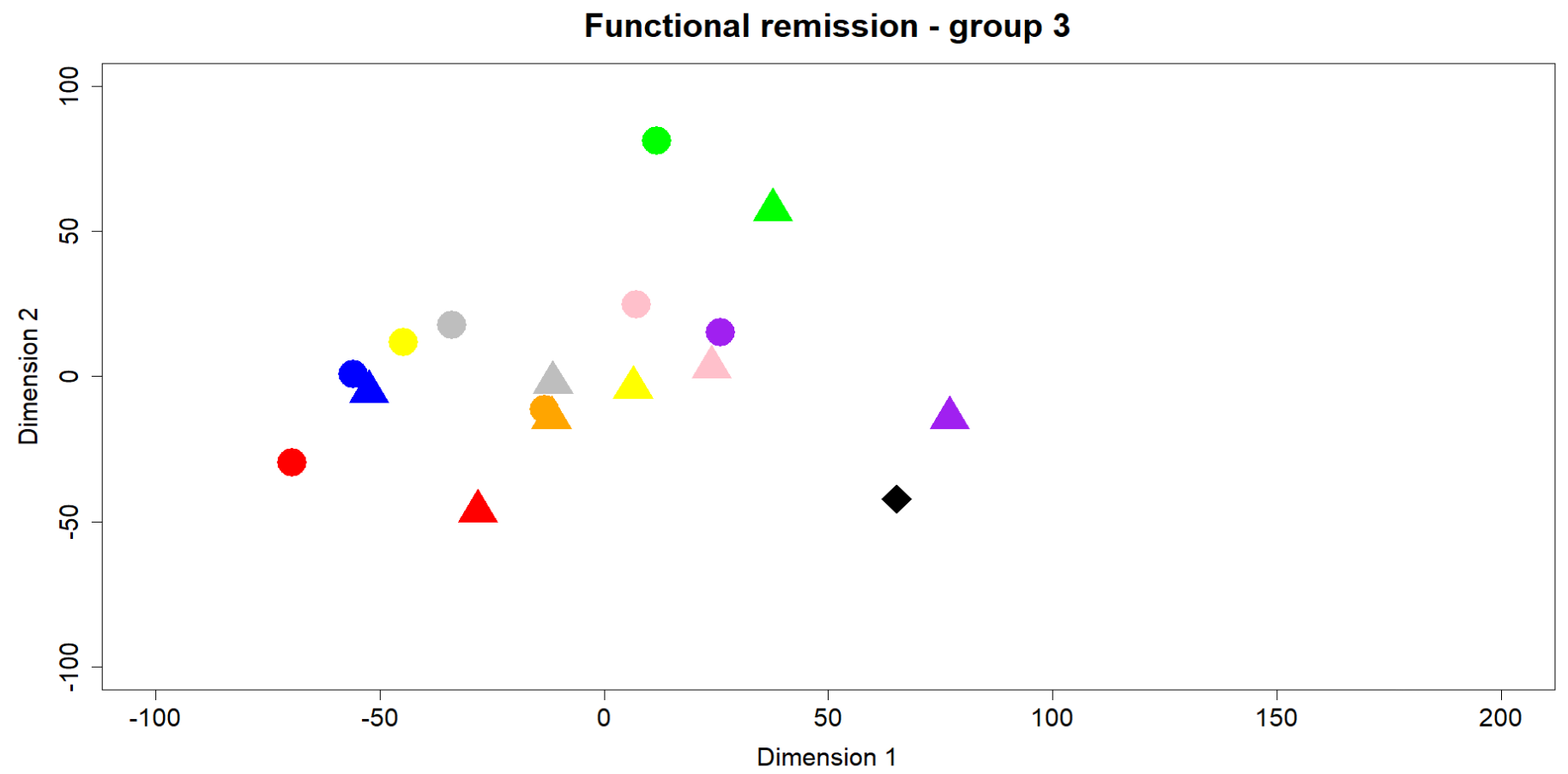

**Supplement 6 -** **Changes in predictions by psychiatrists post-MLM**

|  | Symptomatic remission^1^ | Functional remission^2^ |
| --- | --- | --- |
| Change prediction post-MLM *– absolute* | 25.6% (122/476) | 26.3% (127/482) |
| Mean proportion of changed predictions | 25.8% (range 0% to 77%) | 26.7% (range 5% to 77%) |
| Mean abolute % of change in prognosis prediction | 16.3% (range 0% to 28.5%) | 21.3% (range 6% to 47%). |
| Change in correct direction | 11.6% (55/476) | 15.6% (75/482) |
| Change in wrong direction | 14.1% (67/476) | 10.8% (52/482) |
| Cases with at least one change | 75.8% (50/66) | 72.7% (48/66) |
| Change prediction post-MLM*– dichotomized* | 9.2% (44/476) | 8.7% (42/482) |
| Change in correct direction | 4.2% (20/476) | 4.4% (21/482) |
| Change in wrong direction | 5.0% (24/476) | 4.4% (21/482) |
| Cases with at least one change | 39.4% (26/66) | 37.9% (25/66) |

^1^ For 52 predictions spread over 11 cases there was no ML-prediction available for participants do to a technical error

^2^ For 46 predictions spread over 9 cases there was no ML-prediction available for participants do to a technical error

**Supplement 7 - Visualisation of accuracy of predictions pre- and post-MLM**

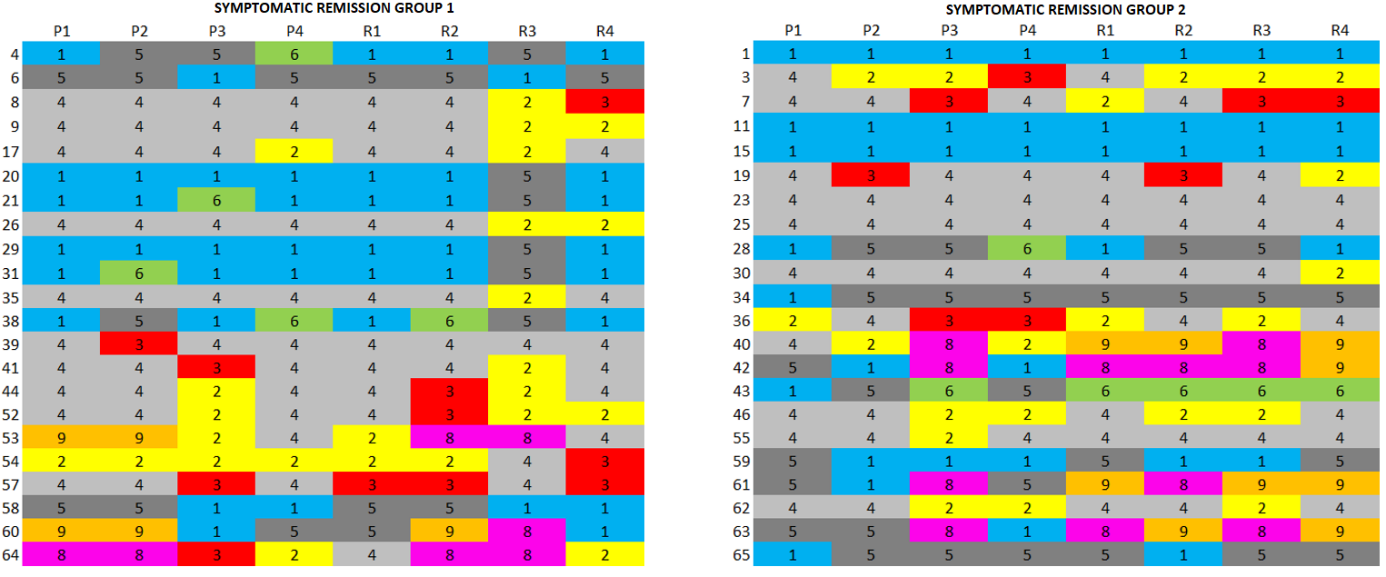

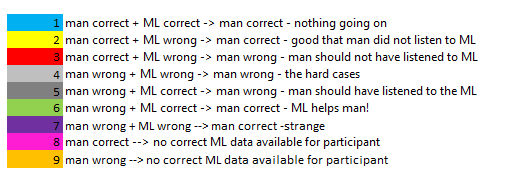

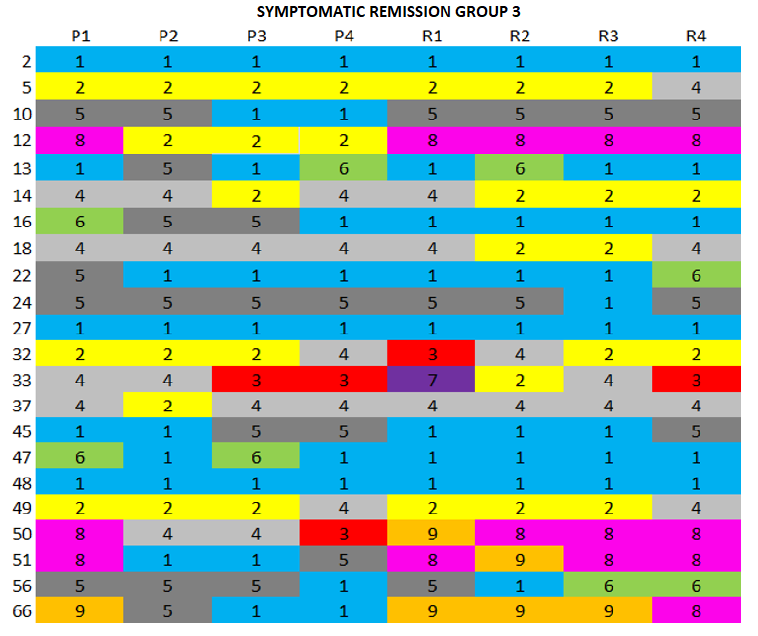

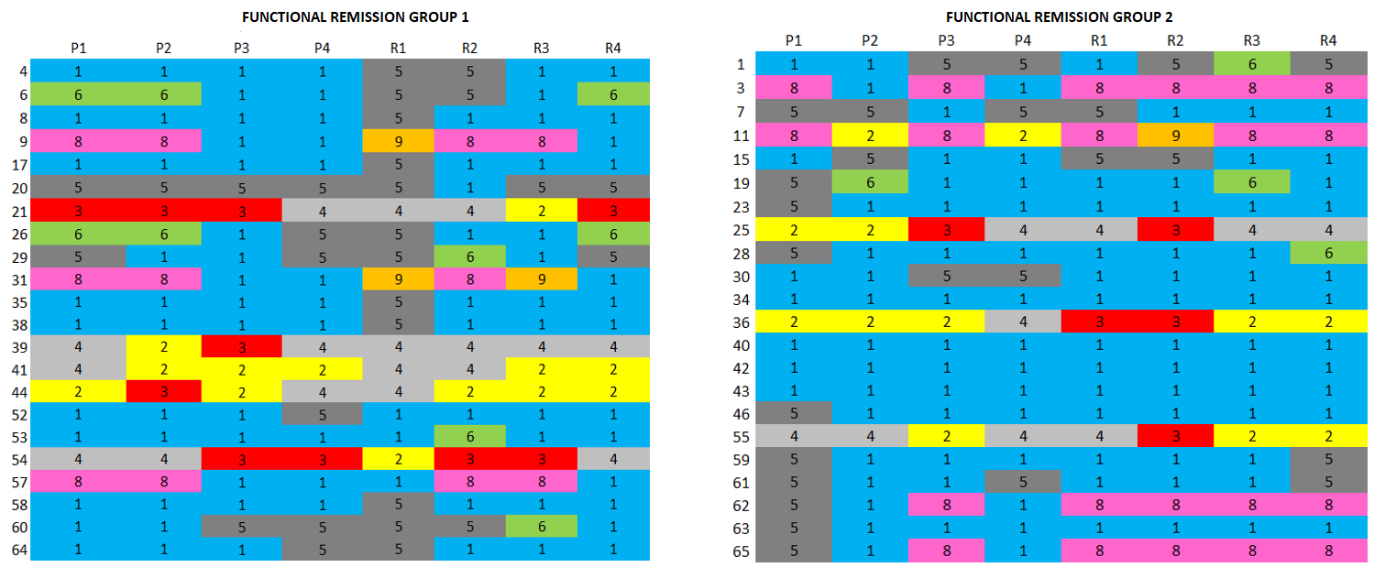

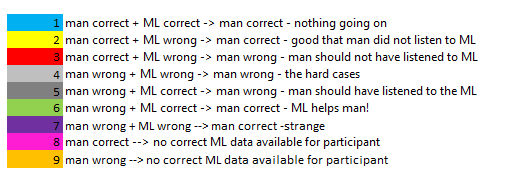

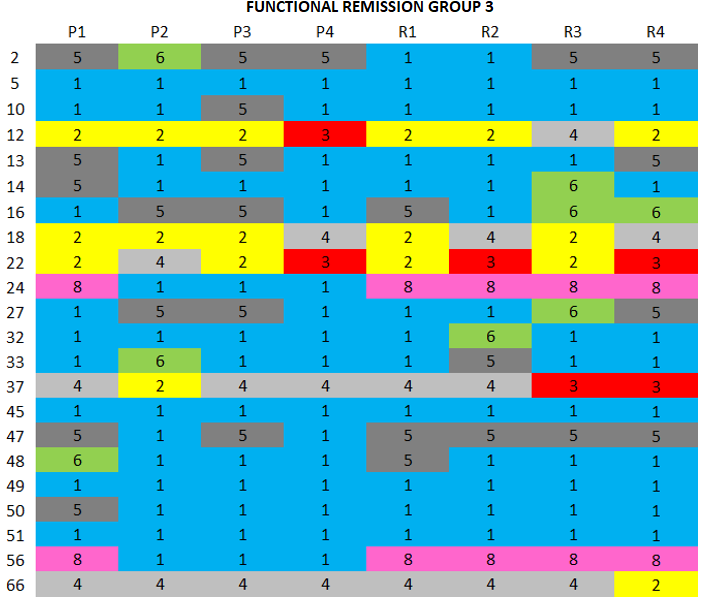

**Supplement 8 - Relative similarity between cases based on patient characteristics**

The following baseline patient information* was used:

Characteristics:

- *Demographic:* age (con), sex (m/f), occupation (y/n), highest level of education (cat), living alone (yes/no).
- *Diagnostic:* DSM-IV classification (Schizophrenia / Schizoaffective disorder / Schizophreniform disorder), current treatment setting (Inpatient / Outpatient / Day Care / Other)

Measurements:

- Calgary Depression Scale for Schizophrenia (CDSS): total score (con)
- Subjective well-being under Neuroleptic Treatment Scale (SWN-K): total score (con)
- Mini International Neuropsychiatric Interview (MINI): mood disorder (y/n), anxiety disorder (y/n), substance abuse/dependence (y/n)
- Positive And Negative Syndrome Scale (PANSS): positive (con), negative (con), general (con), total score (con)
- Personal and Social Performance Scale (PSP): domain A (con), domain B (con), domain C (con), domain D (con), total score (con)
- Clinical Global Impression scale (CGI-severity): total score (con)

*All input features were standardised for use in the t-sne plot

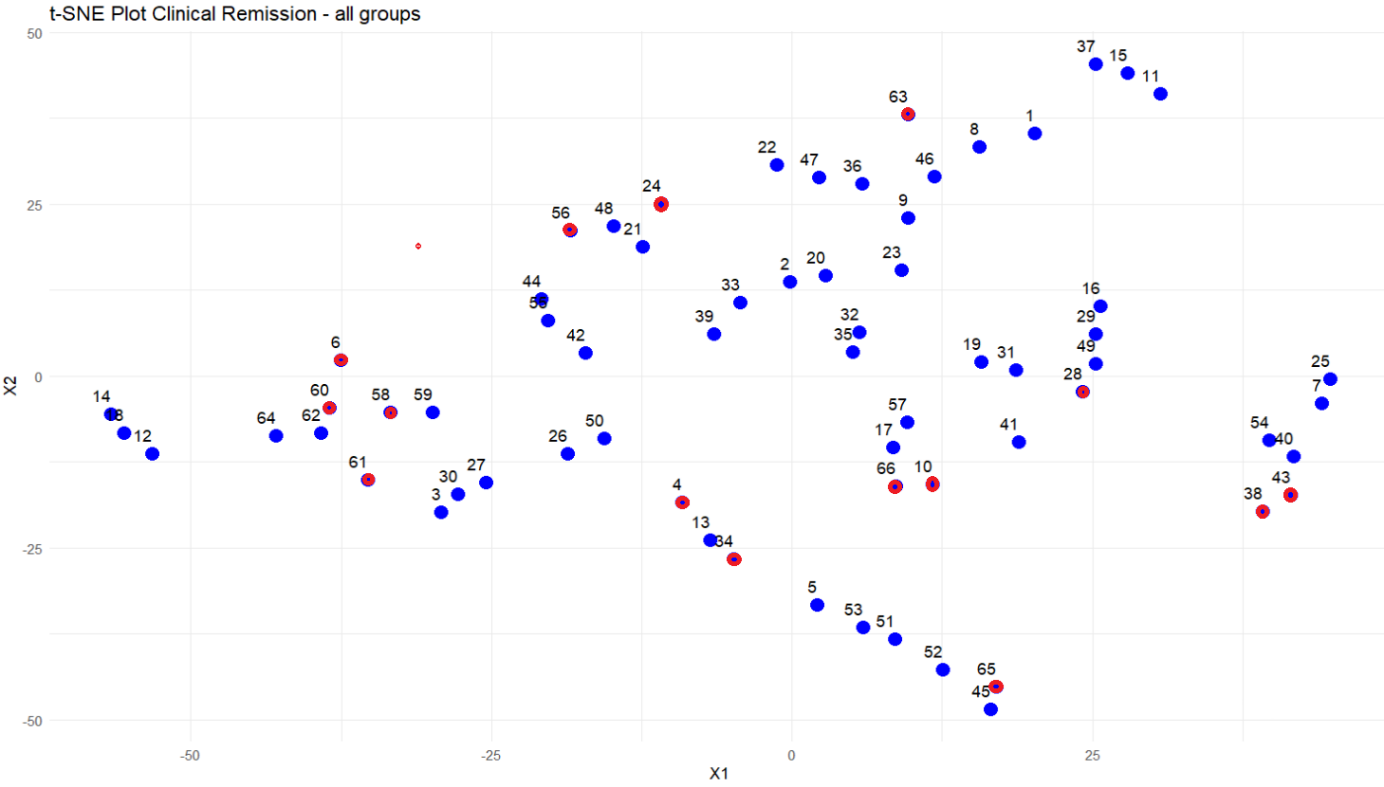

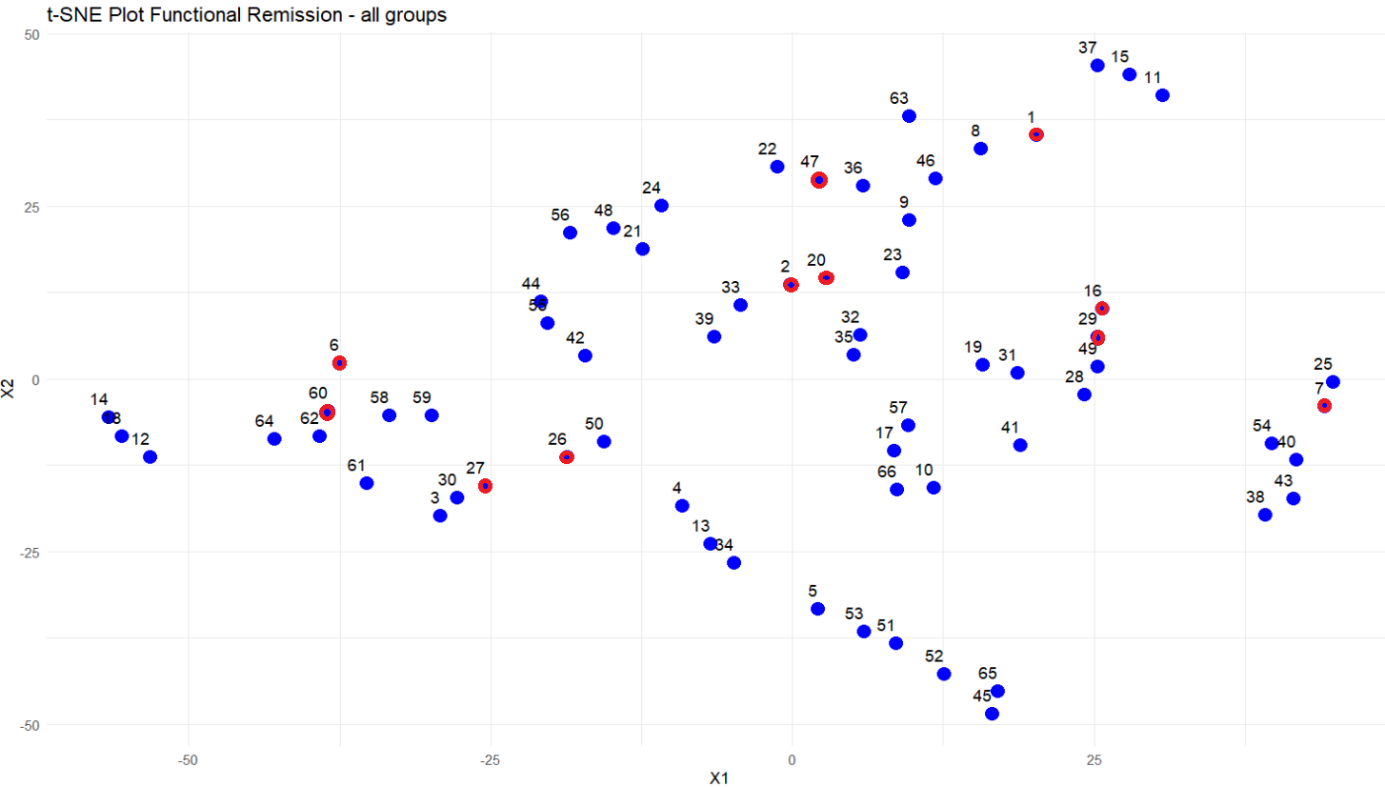

In the t-SNE plot, each case is represented by a data point. The position of the data points relative to each other is determined based on the case characteristics. Cases with more similarities are closer together than those that differ more. Red data points represent the hard cases; the cases that the MLM predicted correctly, while fewer than four psychiatrists did so correctly. The red points are not neatly grouped, meaning they cannot be identified based on case characteristics.
